# Contrasting genetic predisposition and diagnosis in psychiatric disorders: a multi-omic single-nucleus analysis of the human orbitofrontal cortex

**DOI:** 10.1101/2024.04.24.24306179

**Authors:** Nathalie Gerstner, Anna S. Fröhlich, Natalie Matosin, Miriam Gagliardi, Cristiana Cruceanu, Maik Ködel, Monika Rex-Haffner, Xinming Tu, Sara Mostafavi, Michael J. Ziller, Elisabeth B. Binder, Janine Knauer-Arloth

**Affiliations:** Max Planck Institute of Psychiatry, Department Genes and Environment, Munich, Germany; International Max Planck Research School for Translational Psychiatry, Munich, Germany; Institute of Computational Biology, Helmholtz Zentrum München, Neuherberg, Germany; School of Medical Sciences, Faculty of Medicine and Health, University of Sydney, Sydney, Australia; Department of Psychiatry, University of Münster, Münster, Germany; Department of Physiology and Pharmacology, Karolinska Institutet, Stockholm, Sweden; Paul G. Allen School of Computer Science and Engineering, University of Washington, Seattle, WA, USA; Department of Psychiatry and Behavioral Sciences, Emory University School of Medicine, Atlanta, USA

**Keywords:** single-nuclei sequencing, transcriptomic and epigenomic profiling, (regulation of) gene expression, chromatin accessibility, human, postmortem brain, (orbitofrontal) cortex, cell type level/specificity, (severe) psychiatric disorders, genetic risk, multi-omics, large-scale

## Abstract

Psychiatric disorders like schizophrenia, bipolar disorder, and major depressive disorder exhibit significant genetic and clinical overlap. However, their molecular architecture remains elusive due to their polygenic nature and complex brain cell interactions. Here, we integrated clinical data with genetic susceptibility to investigate gene expression and chromatin accessibility in the orbitofrontal cortex of 92 postmortem human brain samples at the single-cell level. Through single-nucleus (sn) RNA-seq and snATAC-seq, we analyzed approximately 800,000 and 400,000 nuclei, respectively. We observed cell type-specific dysregulation related to clinical diagnosis and genetic risk across cortical cell types. Dysregulation in gene expression and chromatin accessibility associated with diagnosis was pronounced in excitatory neurons. Conversely, genetic risk predominantly impacted glial and endothelial cells. Notably, *INO80E* and *HCN2* genes exhibited dysregulation in excitatory neurons superficial layers 2/3 influenced by schizophrenia polygenic risk. This study unveils the complex genetic and epigenetic landscape of psychiatric disorders, emphasizing the importance of cell type-specific analyses in understanding their pathogenesis and contrasting genetic predisposition with clinical diagnosis.

## 1. Introduction

Psychiatric disorders, including major depressive disorder (MDD), bipolar disorder and schizophrenia, have a strong impact on an individual’s quality of life, pose a substantial economic burden, and their most devastating outcome is suicide^1,2^. These disorders not only display overlapping symptoms^3^, but also share a common genetic architecture^4,5^. Genetic correlation analyses have unveiled distinct interconnected clusters among these disorders, indicating their interconnected nature and underscoring the genetic overlap between mood and psychotic disorders^5^. This shared genetic architecture has been the focus of extensive research^6–10^.

Genome-wide association studies (GWAS) have significantly advanced our understanding of the genetic architecture of psychiatric disorders, uncovering numerous significant genetic variants^5,11–13^. Polygenic risk scores (PRS) have emerged as a pivotal tool for capturing the cumulative genetic risk for a particular trait^14^, emphasizing the multi-genic nature of the etiology of psychiatric disorders. The application of PRS has facilitated a deeper understanding of the relationship between genetic risk and various genomic layers^15^, such as gene expression and chromatin accessibility, to understand the full spectrum of psychiatric disorders.

In this context, the role of gene expression studies is particularly relevant. Although previous research has identified numerous genes associated with disorders such as schizophrenia, the direction of effects and overlap with GWAS findings often vary, indicating a complex relationship between gene expression changes and genetic susceptibility^16,17^. Transcriptome-wide association studies (TWAS), expression quantitative trait loci (eQTL) and eQTScore (association analyses between PRS and gene expression) analyses have further bridged the gap between GWAS findings and gene expression data^9,16,18,19^. These integrative approaches provide insights into how the identified GWAS variants can influence gene expression, thereby contributing to the pathophysiology and a more nuanced understanding of psychiatric disorders^20^.

Given that most GWAS variants associated with psychiatric disorders are located in non-coding regulatory elements^5,21^, there is an increased focus on epigenetic studies, which can provide context regarding the intricate relationship between genetics, gene regulation and environmental factors. In this regard Bryois et al. investigated the link between schizophrenia and chromatin accessibility in the prefrontal cortex, identifying their ATAC-seq data as strongly associated with common GWAS variants for schizophrenia^21^. Additionally, Hauberg et al. observed considerable variability in chromatin accessibility across cell types in different cortical regions, revealing that such diversity may obscure cell type-specific effects in aggregate studies, thereby underscoring the intricate complexities of epigenetic regulation^22^. The etiology of psychiatric disorders is notably complex, involving diverse molecular, cellular, and structural alterations across various regions of the human brain, such as the prefrontal cortex, which plays a crucial role in higher cognitive functions and has been linked to various psychiatric conditions^9,11–13,16,23–26^. Structural and functional abnormalities in areas like the orbitofrontal cortex (OFC), a key component of the ventral prefrontal cortex, have been widely reported in many psychiatric disorders^27^. Brodmann area 11, a subregion of the OFC, has shown reduced grey matter volume in schizophrenia patients^28^ and dysregulation of gene expression and DNA methylation in depressed and suicidal patients^29^.

The emergence of single-cell sequencing technologies in recent years has revolutionized our ability to conduct high-resolution studies of various tissues at the level of individual cell types^30,31^. These technologies have enabled the creation of single-cell transcriptomic and epigenomic atlases of the human brain, uncovering hundreds of distinct cell types and even thousands of cellular subtypes within millions of cells across different brain regions^32,33^. Such advancements have greatly enhanced our understanding of the cellular specificity of psychiatric disorders, moving from bulk analyses to the more granular single-cell resolution^26,34–36^.

Our study aims to explore the molecular landscape of the orbitofrontal cortex in psychiatric disorders, using postmortem samples from patients and controls. By examining differential gene expression and chromatin accessibility at the level of single cells, we uncovered key pathways and functions altered across various cortical cell types and how these changes relate to both genetic predisposition and clinical diagnosis. These findings provide new insights into the molecular underpinnings of psychiatric disorders in specific cortical cell types, illustrating how genetic risk factors translate into clinical symptoms and may inform more targeted diagnostics and therapeutic strategies.

## 2. Results

### 2.1. Single-nucleus multi-omic profiling identifies distinct cell types in the human orbitofrontal cortex

To unravel cell type-specific molecular alterations in psychiatric disorders within the orbitofrontal cortex (OFC), we analyzed postmortem brain samples (Brodmann Area 11) using single-nucleus (sn) RNA-seq and ATAC-seq, complemented by genotype information, demographic and clinical variables (Figure 1a). Our cohort was composed of 92 donors, including 35 controls and 57 cases (n_schizophrenia_=38, n_schizoaffective_=7, n_MDD_=7, n_bipolar_=5). Case and control groups were matched for sex, age, postmortem interval and brain pH, see Table S1. Following stringent quality control measures, we obtained high-quality transcriptomic data from 787,046 nuclei, averaging 9,046 nuclei per donor (range 3,895-15,693, Table S2). Each nucleus had a median of 3,887 UMIs, detecting a median of 2,205 genes. Additionally, chromatin accessibility data were acquired for 399,439 nuclei, averaging 4,438 nuclei per donor (range 982-8,707, Table S2) with a median of 7,071 ATAC-seq fragments per nucleus. SnRNA-seq and snATAC-seq data enabled the comprehensive profiling of all major cortical cell types, including excitatory and inhibitory neurons across different cortical layers, endothelial cells, and glial subtypes, like astrocytes, microglia, oligodendrocytes, and oligodendrocyte precursor cells (OPCs). We successfully identified 19 distinct cell types in snRNAs-seq data, with 15 of these also present in the snATAC-seq data (Figure 1b-e). This identification aligns well with the expected diversity of cell types in the human brain and demonstrates the robustness of our method. While the number of nuclei per cell type exhibited heterogeneity, both among different cell types and between snRNA-seq and snATAC-seq datasets (Figure 1e), a high median Pearson correlation coefficient of 0.86 was observed between the cell type proportions of RNA-seq and ATAC-seq data across donors (Supp. Figure 1a). Cell type proportions differed significantly (FDR ≤ 0.05) between the RNA-seq and ATAC-seq modalities for all cell types (Figure 1f, Table S3). Conversely and in agreement with previous research findings^26^, no significant difference in cell type proportions between cases and controls within each data modality were observed (Supp. Figure 1b-c).

**Figure 1.**
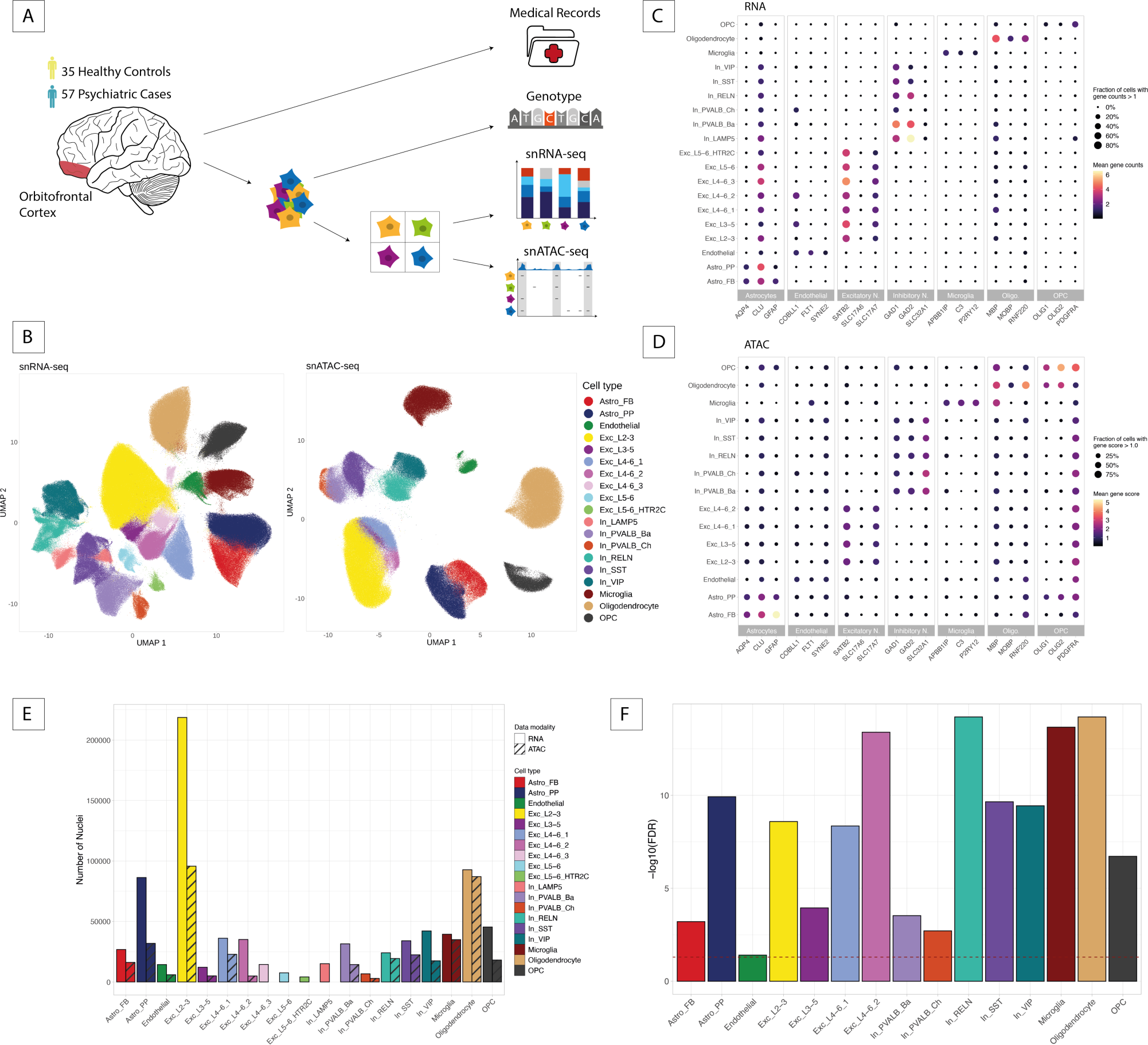
Single-nucleus transcriptomic and epigenomic profiling in the orbitofrontal cortex. (A) Schematic representation of experimental procedures and data modalities. Nuclei were extracted from the orbitofrontal cortex of 57 cases and 35 controls. Single-nucleus (sn) RNA-seq and ATACseq data was integrated with genotype data and medical records. (B) UMAP representations of snRNA-seq (∼800,000 nuclei) and snATAC-seq (∼400,000 nuclei) data colored by the assigned cell type labels. 19 cell types were assigned to the snRNA-seq data and 15 to the snATAC-seq data. (C) Dotplot showing the gene counts in snRNA-seq data of representative marker genes, grouped by major cell types. Color indicates the mean gene counts and size of dots represents the fraction of nuclei with a gene count > 1. (D) Dotplot showing the gene score levels in snATAC-seq data of representative marker genes, grouped by major cell types. Color indicates the mean gene score level and size of dots represents the fraction of nuclei with a gene score > 1.0. (E) Number of nuclei obtained per cell type following quality control colored by cell type. Data modality is indicated by hatching. (F) Significance of differences in cell type proportions between snRNA-seq and snATAC-seq data. Height of the bar represents -log10-transformed FDR values of a two-sided Wilcoxon signed-rank test and the dashed red line corresponds to the FDR cutoff of 0.05.

### 2.2. Cell type-specific alterations in psychiatric disorders: distinct patterns in differential gene expression and chromatin accessibility

As there were no significant differences in cell type proportions between cases and controls within each data modality, we moved our focus on more detailed molecular analyses. To investigate transcriptional alterations associated with psychiatric disorders, we conducted differential expression analyses, contrasting cases (n=57) and controls (n=35) within each cell type (n=19). The extent of significantly differentially expressed (DE, FDR ≤ 0.1) genes varied greatly across cell types, ranging from 0 to 481 (Figure 2a, Table S5). Notably, a high abundance of DE genes was observed within multiple subtypes of excitatory neurons, which also exhibited the most pronounced log_2_-transformed fold changes (FC=[-0.35,0.38], see Figure 2b). More than 50% of DE hits were uniquely dysregulated in one cell type only (Figure 2a), highlighting the distinct transcriptional signatures across cell types. However, the cell types displaying the greatest number of DE genes also possess the highest nuclei count and the largest number of genes evaluated for DE, suggesting a proportional relationship between these variables (see Supp. Figure 3b-c, Supp. Information).

**Figure 2.**
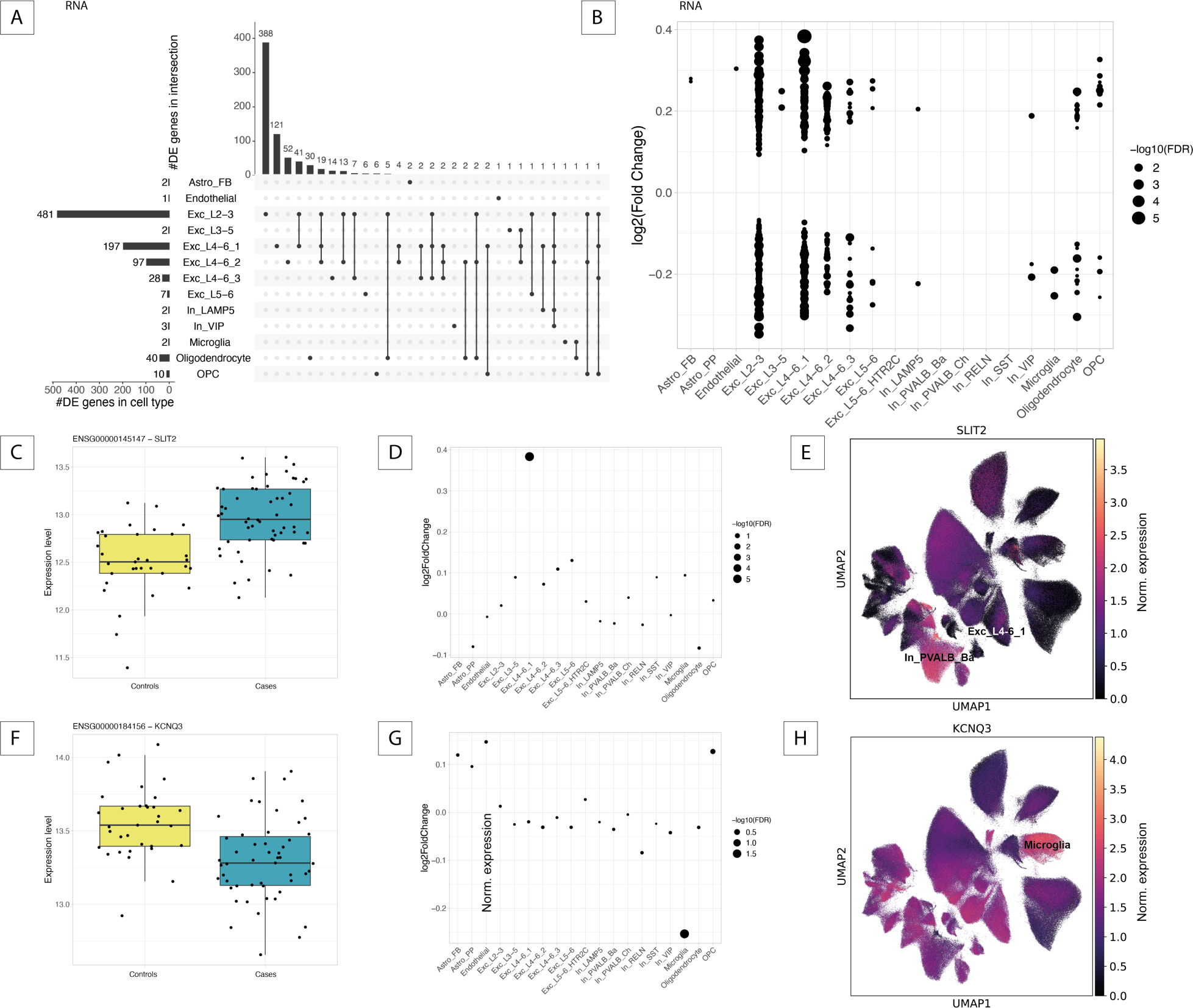
Transcriptional alterations between psychiatric cases and controls. (A) UpSet plot showing the number of differentially expressed (DE) genes (FDR ≤ 0.1) per cell type (left) and the overlap of DE genes between cell types (right). (B) Dotplot of DE genes with log2-fold change on the y-axis and -log10-transformed FDR values represented by dot size. (C,F) Boxplot of normalized gene expression level for *SLIT2* (C) and *KCNQ3* (F) in controls and cases. (D,G) Dotplot of log2-fold changes of *SLIT2* (D) and *KCNQ3* (G) in each cell type. Dot size represents -log10-transformed FDR values. (E,H) UMAP representation of single-nucleus RNA-seq data colored by normalized expression of *SLIT2* (E) and *KCNQ3* (H).

Notably, *Slit Guidance Ligand 2* (*SLIT2)* on chromosome 4 and *Potassium Voltage-Gated Channel Subfamily Q Member 3* (*KCNQ3*) on chromosome 8 demonstrated unique regulatory patterns (Figure 2c-h). *SLIT2* displayed the highest upregulation (FC=0.38, lowest FDR=1.38×10^-6^) in excitatory neurons of layers 4 to 6, cluster 1 (Exc_L4-6_1, Figure 2c-d), despite not exhibiting the highest expression in this cell type (mean exp=0.32, compared to 1.66 in basket cells [In_PVALB_Ba], Figure 2e). While *SLIT2* is known for its role in axon guidance and has been implicated in depression and anxiety behaviors in mice^72^, its specific dysregulation in the human cortex is novel. *KCNQ3* was uniquely downregulated in microglia (FC=-0.25, FDR=0.02), a contrast to its FCs in other cell types (FCs>-0.05, Figure 2f-g). Exhibiting the highest expression in microglia (mean exp=1.75, Figure 2h) and previously linked to bipolar disorder^73^, the specific downregulation of *KCNQ3* in microglia offers new insights into the cellular mechanisms that might contribute to the pathophysiology of this disorder, particularly in the context of neuroinflammation and microglial function.

To evaluate how our results align with previous studies, we correlated our effect sizes with those reported in a single-cell RNA-seq study of schizophrenia in the prefrontal cortex by Ruzicka et al.^26^, see Methods. Among the various correlations observed between the effect sizes of the two studies, those between corresponding cell types were notably the highest. For instance, astrocytes in our dataset exhibited the strongest correlation with those of the corresponding astrocyte population in Ruzicka et al. (Supp. Figure 3d), indicating a broad consistency with previous findings.

From the DE genes we identified within individual cell types (n=872), only 44% (n=387) exhibited a significant difference in gene expression on the full pseudobulk level, which is the aggregated signal of all cell types. Out of all DE genes identified from the full pseudobulk data (n=511), 57% (n=291) were significant in at least one individual cell type (Supp. Figure 3e-f), which highlights the importance of studying the single-cell level.

To complement findings from our DE analysis, we examined variations in chromatin accessibility between cases and controls across shared 15 cell types. Our focus was on differences in gene scores, a quantitative measure of gene activity influenced by accessible chromatin. Only a small number of significant accessibility alterations (DA, FDR ≤ 0.1) were found in two clusters of excitatory neurons (excitatory neurons layers 2/3 [Exc_L2-3], n=45 and excitatory neurons layers 3 to 5 [Exc_L3-5], n=1) and in astrocytes (fibrous [Astro_FB], n=5 and protoplasmic [Astro_PP], n=4), see Figure 3a and Table S6. Only 5 of the DA genes overlapped with DE genes previously identified in the same cell type. When restricting the DA analysis to DE genes within the respective cell type, we found a subset also demonstrating significant alterations in chromatin accessibility. The maximum number of DE/DA genes were 13 in excitatory neurons in superficial layers 2/3 (Figure 3b, Table S7). Notably, discrepancies in regulation direction between transcriptomic and epigenomic data were noted in 8% of DE/DA genes (2 out of 24 genes, Supp. Figure 4a). Among the 22 genes with congruent regulatory patterns in both datasets, Family BHLH Transcription Factor 4 (*HES4*) in excitatory neuron layers 4 to 6, cluster 1 (Exc_L4-6_1), and Insulin-like growth factor-binding protein 5 (*IGFBP5*) in oligodendrocyte precursor cells (OPCs) exhibit the most pronounced fold changes (Figure 3c-d), but also *SLIT2* in excitatory neurons layers 4 to 6, cluster 1 (Exc_L4-6_1) is a DE/DA gene. These findings, including the discrepancies, imply a complex regulatory landscape, and suggest that additional regulatory mechanisms may influence the gene expression.

**Figure 3.**
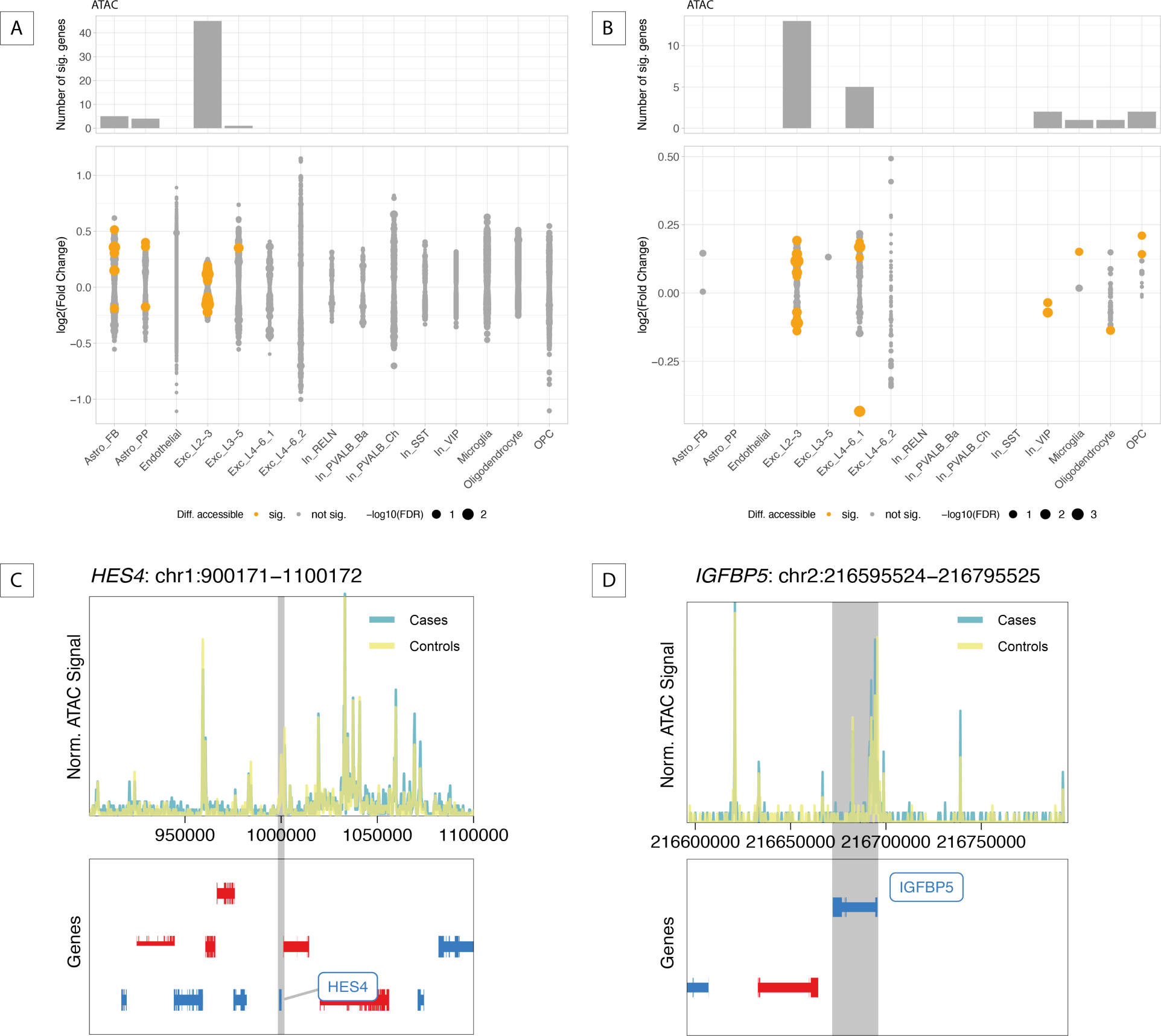
Epigenomic alterations between psychiatric cases and controls. (A-B) Results of differential chromatin accessibility (DA) analysis when testing all genes passing filtering step (A) and only DE genes (B). Barplot on top shows the number of significant DA genes per cell type (FDR ≤ 0.1). Log2-fold changes (FCs) for all tested genes are shown in dotplot with color indicating the DA significance and dot size indicating the -log10-transformed FDR value. No overlap was observed between DA genes in different cell types. (C-D) Genome tracks visualizing normalized ATAC signal in a 100kb window surrounding the gene body of *HES4* (C) and *IGFBP5* (D). *HES4* in excitatory neuron layers 4 to 6, cluster 1 (Exc_L4-6_1), had FDR values of 0.04 (RNA) and 2.21×10^-3^(ATAC) with FCs of -0.28 (RNA) and -0.43 (ATAC). Similarly, *IGFBP5* in oligodendrocyte precursor cells (OPCs) showed FDR values of 1.70×10^-5^ (RNA) and 0.08 (ATAC) with FCs of 0.33 (RNA) and 0.21 (ATAC).

### 2.3. Differential transcriptomic and epigenomic patterns in high vs. low genetic risk donors highlight chromatin accessibility variations

To disentangle the influence of genetic predisposition on gene expression and chromatin accessibility, we utilized polygenic risk scores (PRS) from psychiatric GWAS studies, including cross-disorder phenotype^5^, schizophrenia^74^, MDD^12^ and bipolar disorder^13^, and height^60^ as a non-psychiatric trait (Table S4). Focusing on the extreme PRS groups, matched for confounding variables (see Methods, Supp. Figure 5+6), we found significant DE risk genes in 3 to 10 out of the 19 cell types for each GWAS trait (Figure 4b, Supp. Figure 7a, Table S8). 54 DE risk genes were found in the fibrous astrocytes (Astro_FB) for the cross-disorder phenotype and scattered hits across other cell types (n=18 hits in 5 cell types). Bipolar disorder DE risk genes were detected primarily in excitatory neurons (n=32 out of 35 hits) overlapping partially with the DE genes between cases and controls (n=3 out of 35 hits, Figure 4b, gray dots). Genetic risk for schizophrenia was associated with changes in multiple cell types (n=17 hits in 7 cell types), while fewer MDD risk genes emerged as significant (n=7 hits in 3 cell types). DE risk genes exhibited larger effect sizes than DE genes for clinical diagnoses (median absolute FC_PRS_=[0.29,0.55] vs. median absolute FC_diagnosis_=[0.18,0.30] per cell type, see Supp. Figure 9a). Notably, three DE risk genes were identified across three different cell types for height which are distinct from the DE risk genes for the psychiatric phenotypes.

**Figure 4.**
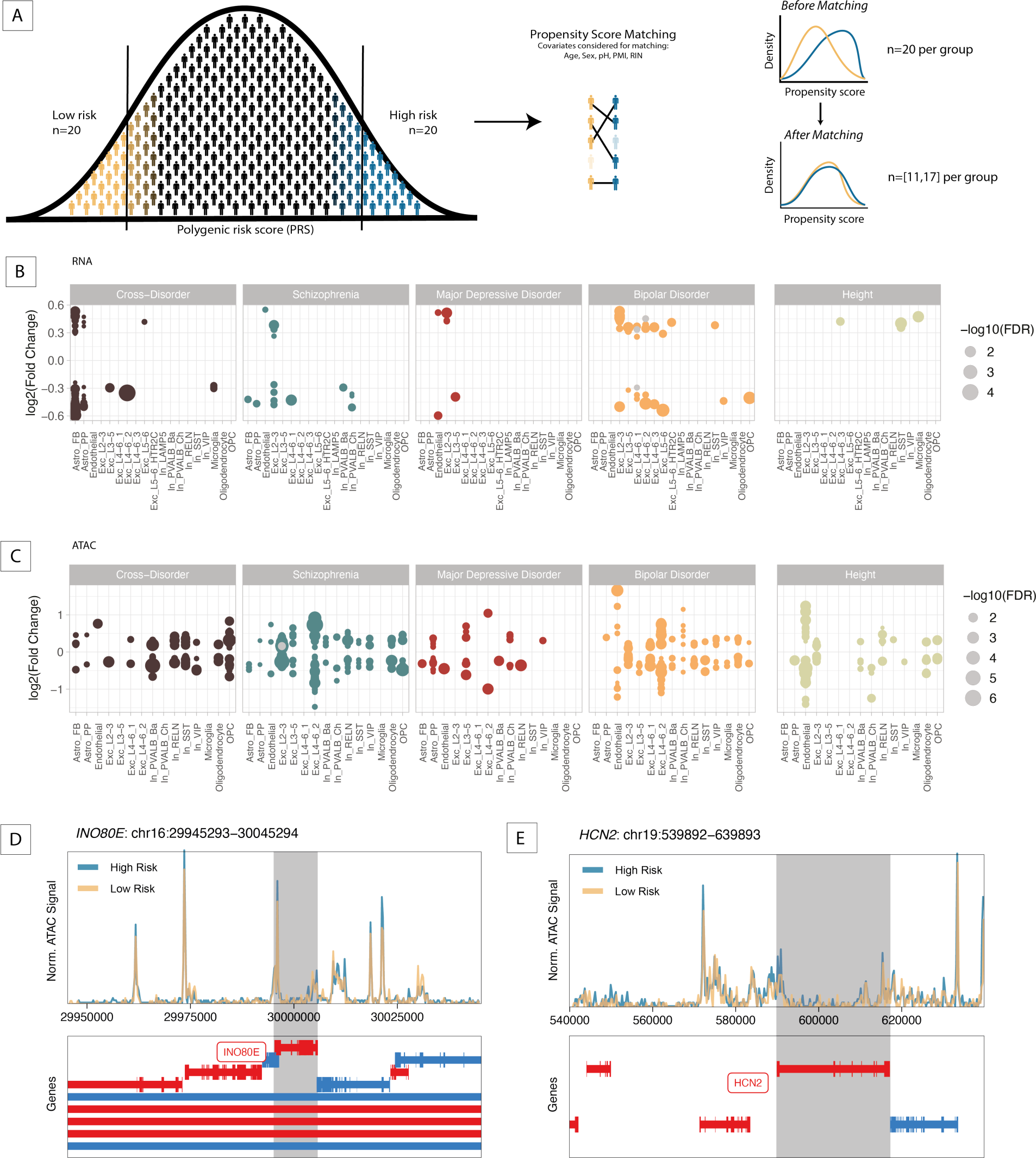
Gene regulatory differences between extreme genetic risk groups. (A) Schematic overview of the definition of extreme genetic risk groups using propensity score matching (see Methods). (B-C) Dotplots of DE (B) and DA (C) genetic risk analyses between extreme genetic risk groups based on 5 different GWAS studies. Each dot represents a significant genetic risk gene (FDR ≤ 0.1). Color indicates the GWAS study, while dot size represents -log10-transformed FDR values. Gray dots in (B) represent DE genetic risk genes that are also DE genes between cases and controls in the same cell type, while gray dots in (C) represent DA genetic risk genes that are also DE risk genes in the same cell type. (D-E) Genome tracks visualizing normalized ATAC signal in a 100kb window surrounding the gene body of *INO80E* (D) and *HCN2* (E).

When investigating DA between extreme PRS groups (DA risk genes), we identified 6,418 DA risk genes across cell types and phenotypes (Figure 4c, Supp. Figure 8a, Table S9), contrasting with 141 DE risk genes. These genes were primarily enriched in excitatory neurons layers 2/3 (Exc_L2-3, n=5,645 DA risk genes). Also DA risk genes exhibited larger effect sizes than DA genes for clinical diagnoses (median absolute FC_PRS_=[0.15,0.74] vs. median absolute FC_diagnosis_=[0.12,0.35] per cell type, see Supp. Figure 9b). Notably, despite identifying DA risk genes for height, only one overlapped with bipolar disorder DA risk genes. The overlap between DA and DE risk genes was minimal with only two genes (Figure 4c, gray dots), *hyperpolarization-activated cyclic nucleotide-channel* (*HCN2)* and *IN080 complex subunit E (INO80E*), being both DE (FCs=0.36 and 0.26, FDR=0.06 and 0.09 respectively) and DA (FCs=0.14 and 0.16, FDR=0.05 and 0.03 respectively) for schizophrenia risk in excitatory neurons layers 2/3 (Exc_L2-3). Genomic tracks surrounding *HCN2* and *INO80E* illustrate different ATAC coverage for the high and low schizophrenia risk groups (Figure 4d-e). In bulk GTEX data, *HCN2* is mostly expressed in the heart and the nervous system^75^ (Supp. Figure 8f) and contributes to pacemaker currents^76^, while *INO80E* is expressed across all tissues^75^ (Supp. Figure 8f). *INO80E* is involved in ATP-dependent chromatin remodeling, DNA replication and repair^77^.

In the analysis of the overlap between diagnosis-related genes and genetic risk genes, our findings revealed a distinct cellular specificity. The molecular response in neurons was influenced by both diagnosis-related genes (DE or DA genes) and genes associated with genetic risk (DE or DA risk genes). In contrast, glial cells predominantly exhibited molecular alterations linked to genetic risk factors. Specifically, 81% of gene alterations in OPCs, 76% in microglia, and over 90% in both fibrous (Astro_FB) and protoplasmic astrocytes (Astro_PP) were linked to genetic risk rather than disease status. Endothelial cells were also primarily influenced by genetics, with 95% of changes tied to genetic risk (Supp. Figure 10).

### 2.4. Disease-relevant pathway enrichment in microglia is uncovered by transcriptomic profiling

To explore the biological processes and functions affected by genes differentially regulated due to diagnosis or genetic predisposition within different cell types, we conducted KEGG pathway enrichments for each cell type in the 250 most up- and downregulated DE and DA genes and DE and DA risk genes (see Methods). For the top DE genes between cases and controls, downregulated genes in microglia were distinctively enriched for pathways like long-term depression (FDR=0.04) and cell-cell interaction mechanisms, such as focal adhesion (FDR=0.04), setting them apart from other cell types (Figure 5). Furthermore, top DE genes highlighted distinct pathways in the nervous and endocrine systems (e.g. various synapses or endocannabinoid signaling) enriched for downregulated genes in fibrous astrocytes (Astro_FB), chandelier cells (In_PVALB_Ch), and microglia. Pathways related to neurodegenerative diseases and oxidative phosphorylation showed significant enrichment in both up- and downregulated genes in different cell types. Notably, the Ribosome pathway exhibited significant enrichment, particularly in upregulated genes especially in oligodendrocytes (FDR=5.18×10^-29^), alongside moderate upregulation observed in OPCs (FDR=7.20×10^-7^) and endothelial cells (FDR=6.54×10^-11^). Many pathways enriched in up- and downregulated DE risk genes reflecting genetic risk (Supp. Figure 7c-f) overlap with the pathways enriched in genes altered between cases and controls, while the respective cell types exhibiting the enrichment are often different. For chromatin accessibility alteration between cases and controls, pathway enrichment analysis revealed no significant enrichments for most cell types (Supp. Figure 4b) and for extreme genetic risk groups it revealed only few significant pathways (Supp. Figure 8b-e) which can be attributed to the DA genes’ involvement in separate biological processes rather than shared pathways, and different sizes of background sets.

**Figure 5.**
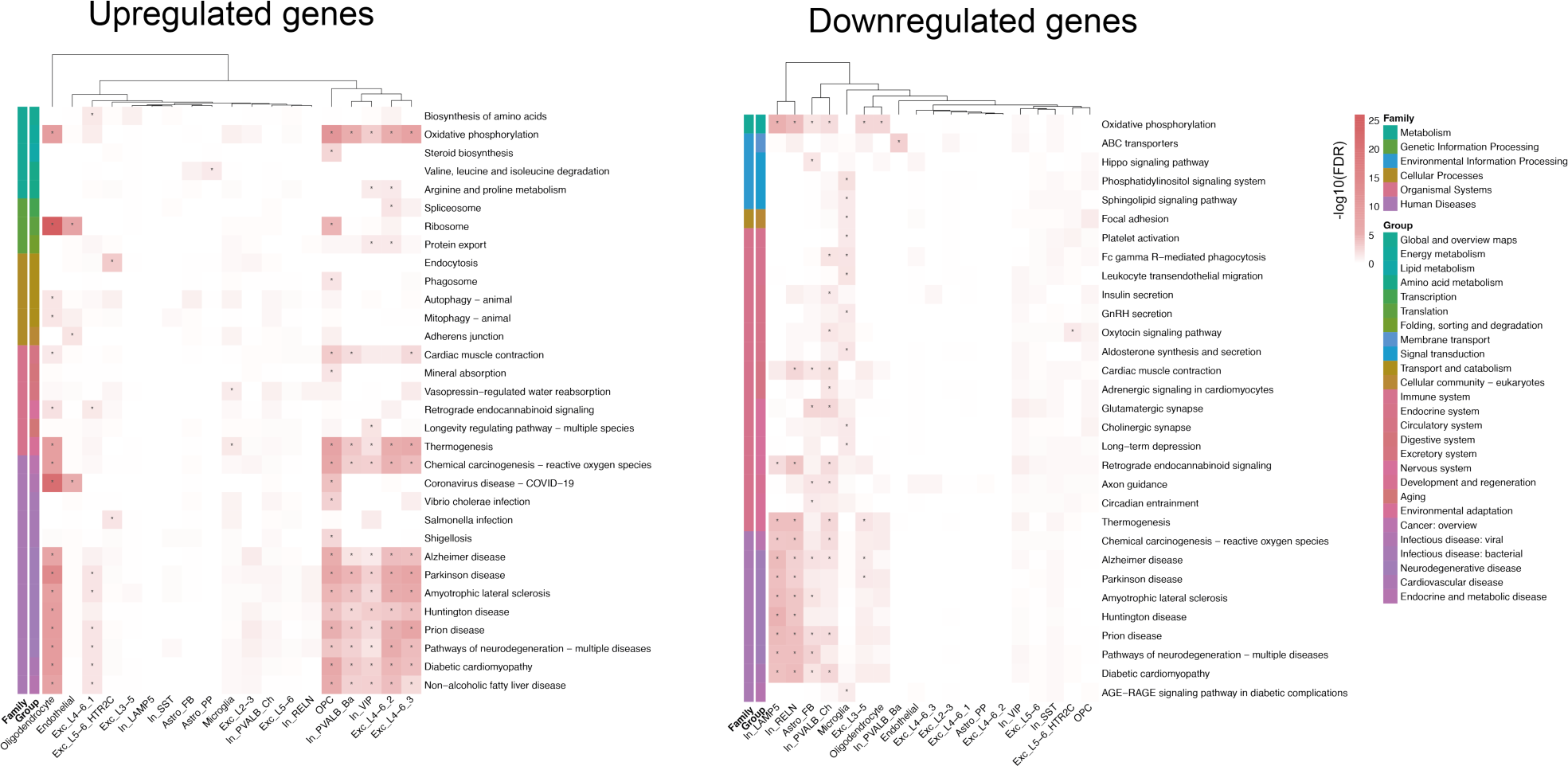
Disease-relevant pathway enrichments. Heatmaps of KEGG pathway enrichment results for the 250 most up- and downregulated DE genes per cell type comparing cases and controls. All pathways significantly enriched in at least one cell type are included in the heatmap, with color representing -log10-transformed FDR values and asterisks indicating significance (FDR ≤ 0.05). Pathway annotations on the left indicate their pathway group and family, and dendrograms visualize k-means clustering of cell types according to enrichment results.

### 2.5. Schizophrenia polygenic risk influences *INO80E* and *HCN2* regulation in excitatory neurons in superficial layers 2/3, independent of diagnosis

The gene *INO80E*, previously linked to schizophrenia through genomic studies including GWAS, transcriptome-wide association analysis, and copy number variation analysis^18,78–81^, emerged as a significant DE and DA risk gene in schizophrenia PRS extreme groups, specifically in excitatory neurons in superficial layers 2/3 (Exc_L2-3, Figures 4c-d, 6a-c), yet showed no association with disease status. We explored its regulatory mechanisms using a correlation-based network that included gene expression, chromatin accessibility, PRSs, and disease status to visualize the multi-omic data used in our study (Figure 6d). The network revealed positive correlations within nodes of the same data modality but negative correlations across different node types. Notably, *INO80E* exhibited differential accessibility in Exc_L2-3 among extreme genetic risk groups for cross-disorder and schizophrenia PRS. However, its correlation with gene expression fell below nominal significance, despite being a significant DE risk hit. Transcription factor motif enrichment analysis in *INO80E’s* promoter region identified significant *KLF4* motif enrichment (Table S10, Figure 6e). Although *KLF4* has been associated with schizophrenia and reported to be downregulated in patients^82^, it was not expressed in Exc_L2-3 in our dataset.

**Figure 6.**
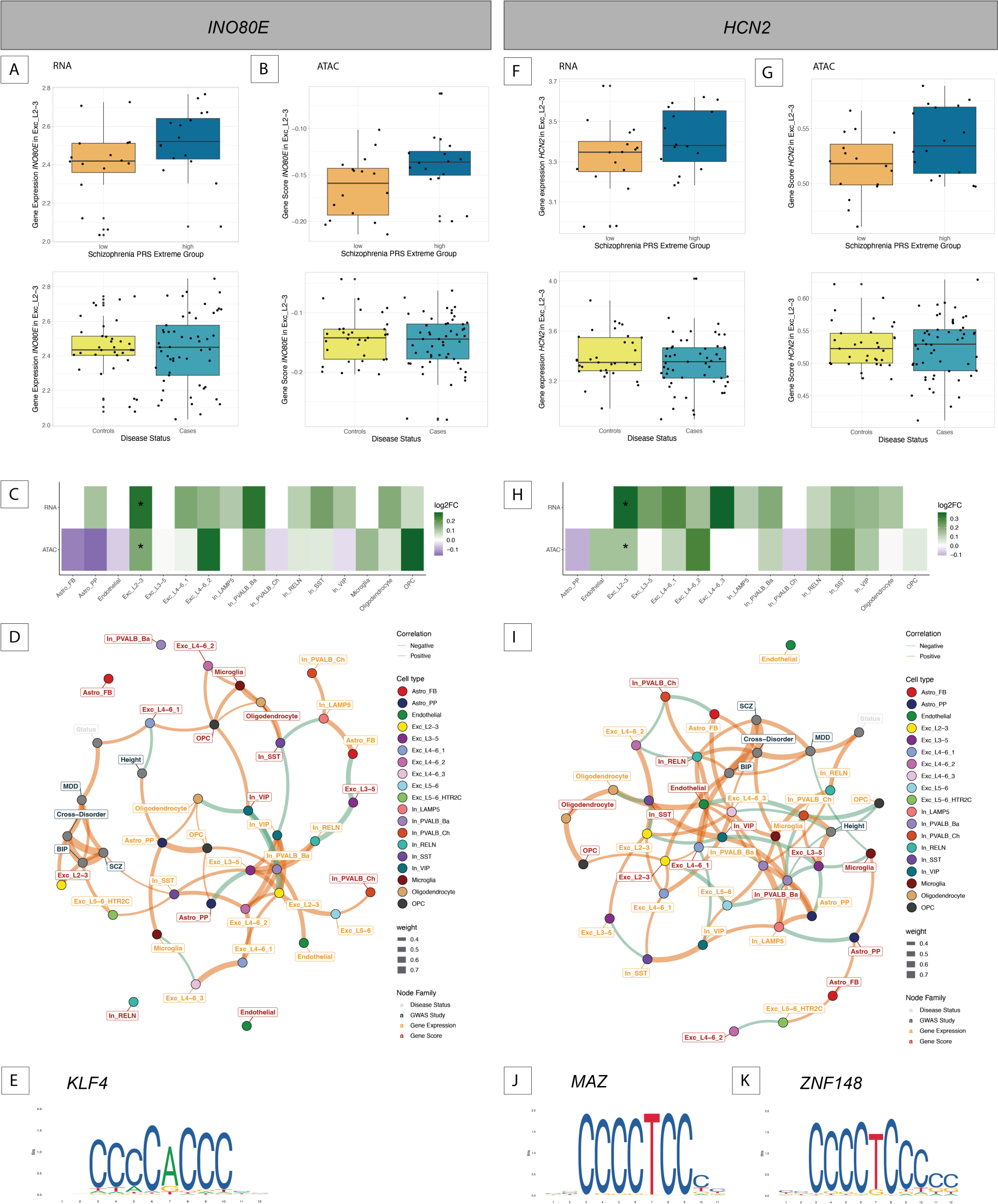
Cell type-specific gene regulation of *INO80E* and *HCN2* related to genetic risk for schizophrenia. (A,B,F,G) Boxplots showing gene expression (A,F) and chromatin accessibility (B,C) of *INO80E* (A,B) and *HCN2* (F,G) for extreme genetic risk groups as well as disease status. (C) Heatmap visualizing the log2-fold changes of *INO80E* from DE and DA genetic risk analysis for schizophrenia. Asterisks indicate significance (FDR ≤ 0.1). (D,I) Correlation-based network for *INO80E* (D) and *HCN2* (I) inferred from PRS for cross-disorder phenotypes, bipolar disorder, MDD, schizophrenia and height, disease status as well as gene expression and chromatin accessibility across cell types. All nominally significant correlations are shown (P ≤ 0.05). Node color indicates the cell type, color of node labels indicates the node family/data modality, edge thickness relates to correlation strength and edge color indicates whether the correlation is positive or negative. (E,J,K) Transcription factor motif for *KLF4* (E), *MAZ* (J), and *ZNF148* (K), as sourced from JASPAR^69^.

A second gene, *HCN2*, coding for a hyperpolarization-activated cation channel crucial in pacemaker activity in the heart and brain^76^, showed differential expression and accessibility (DE and DA risk gene) in Exc_L2-3 among extreme genetic risk groups for schizophrenia PRS (Figure 4c,e, 6f-g), with no significant dysregulation in other cell types (Figure 6h). The correlation-based network analysis for *HCN2* (Figure 6i) indicated more positive correlations between data modalities than the *INO80E* network. Only with the network approach, *HCN2*’s gene scores in Exc_L2-3 were positively correlated with bipolar disorder PRS and its expression in Exc_L2-3 showed positive correlations with other excitatory neuron populations, but negative correlations with VIP and SST interneurons (In_VIP and In_SST, Figure 6i). Transcription factor motif analysis in the *HCN2* promoter identified several significant motifs (Table S10), notably for *MAZ* (Figure 6j) and *ZNF148* (Figure 6k), with these genes being expressed and accessible in Exc_L2-3 and most other cell types.

In summary, the network analysis emphasizes the importance of considering both genetic predisposition and diagnosis, particularly evident in the two genes identified as significant DE and DA genetic risk hits simultaneously, yet not as DE or DA genes. Such networks underscore the complex interplay of transcriptional and epigenetic mechanisms, providing valuable insights that deepen our understanding of schizophrenia.

## 3. Discussion

In this study, we performed a comprehensive single-nucleus analysis on postmortem brain tissue from 92 donors, including 57 with psychiatric diagnoses. This research stands out for its extensive scale, analyzing gene expression in roughly 800,000 nuclei and assessing chromatin accessibility in approximately 400,000 nuclei from the orbitofrontal cortex, marking a significant advancement in nuclei count for a single-cell study in psychiatric research. Our findings revealed crucial differences in gene expression and chromatin accessibility primarily associated with genetic risk rather than diagnosis. Notably, glial cells predominantly showed molecular alterations associated with genetic risk genes, while neurons demonstrated a molecular response influenced by both diagnosis-related and genetic risk genes. Additionally, we identified distinct pathway enrichments in downregulated genes in microglia.

In our study, the majority of genes exhibit differential expression in excitatory neurons among psychiatric cases and controls, consistent with prior literature findings^26^. However, it is essential to consider variations in detection power among cell types when interpreting these findings. Particularly noteworthy are *SLIT2* and *KCNQ3*, previously implicated in psychiatric disorders but without specific cell type associations^72,73^. Our study reveals cell type-specific pathological alterations for these genes. *SLIT2,* exclusively dysregulated in excitatory neurons layers 4 to 6, cluster 1, has been linked to depression- and anxiety-like behaviour in mice^72^, the development of serotonergic and dopaminergic circuits in the forebrain^83^, and shaping vulnerability to suicide attempts^84^. Additionally, *KCNQ3,* was specifically downregulated in microglia, previously associated with reduced gene expression and altered DNA methylation in bipolar disorder^73^, and proposed as a novel target in depression and anhedonia treatment^85^. Our cell type-specific findings expand upon previous bulk tissue studies, offering insights for more in-depth investigations into disease consequences and potential new directions for therapeutic research. For instance, drugs targeting *KCNQ3* expression in microglia could modulate gene expression levels or target specific cellular pathways involved in *KCNQ3* expression to counteract disease pathology.

Our study aimed to investigate the correlation between gene expression and chromatin accessibility to discern if expression changes were influenced by chromatin alterations. Our findings indicate that at the cell type level there is generally a positive correlation between ATAC-seq signals near a gene and its expression (Supp. Figure 2c-d), consistent with prior research^86^. However, deviations exist for many genes, as observed in studies such as that by Sanghi et al., which identified less than 20% correlation for differential promoter elements and gene expression, and less than 5% correlation for non-differential promoter elements in a cancer cohort study^87^. Specifically, out of 872 differentially expressed genes, 867 exhibited changes in gene expression without corresponding alterations in nearby chromatin accessibility. This suggests that transcriptomic changes in psychiatric disorders could occur independently of alterations observed in chromatin accessibility. Moreover, focusing on differentially accessible genes corresponding to differentially expressed genes, we identified a small subset of genes with alterations in both gene expression and chromatin accessibility between psychiatric cases and controls (n=24 out of 872 genes). For instance, *HES4* was consistently downregulated in gene expression and chromatin accessibility in excitatory neurons layers 4 to 6, cluster 1, previously associated with abnormal psychomotor behavior in schizophrenia^88^. Epigenetic dysregulations in *HES4* have also been shown to be related to neuronal development and neurodegeneration in postmortem brains^89^. Similarly, *IGFBP5*, showed upregulation in both gene expression and chromatin accessibility in OPCs, associated with depressive symptoms and cognitive dysfunction in aging^90^. These findings highlight the complex interplay between gene expression and chromatin accessibility in psychiatric disorders. Considering that environmental factors like stress or lifestyle choices, relevant to psychiatric diseases^91^, can influence epigenetic modifications and affect chromatin structure and gene expression^92^, exploring these relationships becomes even more crucial. Further studies employing alternative or more specific types of epigenetic regulation, such as histone modification analysis, may provide deeper insights into the regulatory mechanisms underlying psychiatric disorders.

Building on our findings, we examined the significance of genetic risk factors. Our analysis showed distinct regulatory patterns associated with genetic risk across disorders and specifically in bipolar disorder, MDD and schizophrenia, which were highly different between traits and from those associated with diagnosis. This difference highlights the complex interplay between genetic and epigenetic factors, which are likely influenced by environmental factors in psychiatric disorders^91^. The little overlap of differentially expressed genes associated with genetic risk for psychiatric disorders and differentially expressed genes associated with disease status underscores the importance of investigating genetic risk independently of clinical diagnosis. This distinction becomes crucial given the inherent limitations and variability of diagnosis, which might not always accurately reflect the actual nature or severity of a psychiatric condition^3,93^. Our study contributes to the foundation for future research aimed at integrating these findings into a comprehensive tool that considers both risk factors and diagnoses, not just in isolation but in a synergistic manner for enhanced diagnostic and research applications. By focusing on extreme groups rather than treating genetic risk as a continuous variable, we applied a robust approach that has demonstrated reliability in previous studies^11–14,62^.

Our findings highlight distinct influences of genetic risk and clinical diagnosis on various cell lineages, particularly neuronal and glial populations, with genetic risk genes exhibiting larger effect sizes (Supp. Figure 9). Excitatory neurons, a key neuronal class, exhibited significant alterations influenced by diagnosis as well as genetic risk in both ATAC-seq and RNA-seq data. This aligns with the understanding that neuronal populations are directly involved in the synaptic and circuit-level changes often associated with psychiatric conditions^94^. In contrast, endothelial cells and glial populations, such as astrocytes, OPCs and microglia, were more distinctly influenced by genetic risk factors (76%-97% of the genes are unique DE or DA risk genes). This intriguing result suggests that glial cells, traditionally viewed as support cells, may play a more active role in the genetic predisposition to psychiatric disorders^95^. The differential impact observed across cell lineages suggests that while some may have a genetic predisposition to psychiatric disorders, others are more reactive to disease-associated pathophysiological changes, underscoring the importance of examining neuronal, endothelial, and glial roles in psychiatric research. This distinction was not as evident in previous bulk studies, which often obscured cell type-specific dynamics due to their aggregated nature.

Our study revealed minimal overlap of genes between the diagnostic and genetic risk analyses. However, an interesting aspect is that despite this minimal overlap, the affected pathways often correspond. This observation may stem from the functional convergence of affected genes, besides a potential lack of power to detect more overlapping genes. For instance, we identified a significant enrichment of ribosomal processes in genes upregulated in oligodendrocytes, OPCs, and endothelial cells. This finding aligns with previous research linking ribosomal dysregulation to psychiatric disorders^96,97^. Dysfunction in ribosomal processes could impact key features like protein synthesis and synaptic function in psychiatric conditions^98^. Furthermore, pathways related to neurodegenerative diseases and oxidative phosphorylation were enriched in various cell types, suggesting complex regulation across cell populations. Perturbations of protein synthesis as well as oxidative stress, usually caused by an imbalance of oxidative phosphorylation and the removal of its byproduct, can lead to excitation/inhibition imbalance implicated in the pathophysiology of schizophrenia^97,99^. Specifically, the unique dysregulation pattern observed in genes downregulated in microglia reflects their critical role in brain health^100^, potentially linked to increased inflammation and stress-induced brain changes implicated in psychiatric disorders like schizophrenia^101–103^. The ability of microglia to adapt and switch roles in response to inflammation^104^ may underlie this observed unique dysregulation pattern, reflecting their complex and multifaceted roles in the intricate relationship between microglia, inflammation, and psychiatric conditions.

While our analysis revealed very few differentially accessible genes between cases and controls, we noticed a significantly higher number of differentially accessible genes compared to differentially expressed genes associated with genetic risk for psychiatric disorders (6,418 vs. 141, Supp. Figure 7a+8a). The only genes observed as significant in both differential expression and accessibility analysis for genetic risk are *INO80E* and *HCN2* in excitatory neurons layers 2/3 for schizophrenia. *INO80E* has been highlighted before as a promising drug target as it is a GWAS, TWAS and CNV hit for schizophrenia^78^, while *HCN2* has been identified as differentially methylated in the prefrontal cortex and hippocampus in schizophrenic patients^105,106^ and its knockdown leads to antidepressant behavior in rodents^107,108^. Taken together, these results suggest that chromatin accessibility alterations play a more prominent role in the genetic basis of psychiatric disorders compared to changes in gene expression levels. Chromatin accessibility may represent an earlier or more fundamental level of genetic regulation, influencing a gene’s potential for expression before actual changes in gene expression occur, as it is also the case for developmental processes in the cortex^109^. This implies that chromatin accessibility serves as a more sensitive marker for genetic predispositions to psychiatric disorders.

Following our observation of distinct genetic risk mechanisms and diagnosis, and with the ultimate goal of creating a diagnostic tool, we conducted an integrative correlation-based network analysis of the key affected genes, including *INO80E* and *HCN2*. It revealed that correlations tend to be positive within the same data modality (e.g. gene expression, PRS) and negative between different modalities, indicating that gene expression or accessibility differences linked to genetic risk may not align with linear trends in genetic risk scores, though these scores tend to correlate more with chromatin accessibility than gene expression. In exploring regulatory elements of *INO80E* and *HCN2*, our transcription factor motif analysis identified a *KLF4* motif enrichment for *INO80E*. The absence of *KLF4* expression and accessibility in excitatory neurons layers 2/3 in our dataset suggests a potential mismatch in the timing of *KLF4* expression or the involvement of a different, unidentified transcription factor. For *HCN2*, the numerous enriched motifs suggest intricate, cell-context-specific transcriptional regulation.

Our findings highlight the complexities in gene regulation. The inconsistent correlation between gene expression and chromatin accessibility suggests that other regulatory mechanisms, including trans-regulatory elements and additional epigenetic layers like DNA methylation and histone modifications, are at play. These aspects, while not covered in our study, underscore the multifaceted nature of genomic regulation in psychiatric disorders. Our study, with one of the largest single-cell datasets in psychiatric research (n=92 donors and ∼800,000/400,000 nuclei), marks a significant advancement in understanding these disorders. Although the study is substantial, further research is needed to fully capture the genetic and epigenetic diversity in the affected population. Additionally, our cohort’s limitation to individuals of European ancestry highlights a common issue in psychiatric research: the underrepresentation of diverse populations in biosample collections. There is a critical need within the research community to gather more inclusive samples, ensuring findings are applicable across varied ethnic backgrounds.

Subsequent investigations could include the inference of cell type-specific gene regulatory networks, offering a more comprehensive view of the regulatory mechanisms underlying the observed transcriptional and epigenomic alterations. Building on the identified molecular signatures, future research also needs to focus on developing targeted therapies or repurposing existing drugs to specifically address the underlying molecular perturbations associated with psychiatric disorders.

## 4. Methods

### 4.1. Human postmortem brain samples

As previously described^37,38^, ethics approval was obtained from both the Ludwig Maximilians-Universität (22-0523) and the Human Research Ethics Committees at the University of Wollongong (HE2018/351). Donors or their next of kin provided informed consent for brain donation. Utilizing fresh-frozen postmortem tissues of the orbitofrontal cortex (Brodmann area (BA) 11 dissected from the 3rd 8-10mm coronal slice), sourced from the NSW Brain Tissue Resource Centre in Sydney, Australia, we conducted single-nucleus RNA-sequencing (snRNA-seq) and single-nucleus ATAC-sequencing (snATAC-seq). Our study encompassed a cohort of 92 donors. This included 35 psychiatrically healthy controls and 57 cases diagnosed with schizophrenia, schizoaffective disorder (SCA), major depressive disorder (MDD), or bipolar disorder (n=38;7;7;5, respectively). The case and control groups were matched in terms of sex (38% female representation), age (mean ± s.d. = 54.27 ± 13.64), postmortem interval (mean ± s.d. = 33.90 ± 14.82), and brain pH (mean ± s.d. = 6.60 ± 0.24), see Table S1.

### 4.2. Nuclei isolation and single-nucleus RNA and ATAC sequencing

Nuclei were extracted from ∼50 mg frozen postmortem brain tissue (BA11) as previously described^37^. In short, tissue was homogenized using dounce-homogenization in 1 ml nuclei extraction buffer (10 mM Tris-HCl pH 8.1, 0.1 mM EDTA, 0.32 M Sucrose, 3 mM Mg(Ac)_2_, 5 mM CaCl_2_, 0.1% IGEPAL CA-630, 40 U/ml RiboLock RNase-Inhibitor (ThermoScientific)). Next, homogenate was layered onto 1.8 ml of sucrose cushion (10 mM Tris-HCl pH 8.1, 1.8 M Sucrose, 3 mM Mg(Ac)_2_) and ultra-centrifuged at 28,100 rpm at 4°C for 2.5 hours (Thermo Scientific™ Sorvall™ WX+ 471 ultracentrifuge). Using vacuum suction supernatant was removed and nuclei pellet was gently resuspended in 80 μl resuspension buffer (1X PBS, 3 mM Mg(Ac)_2_, 5 mM CaCl_2_, 1% BSA, 40 U/ml RiboLock RNase-Inhibitor). From the same nuclei suspension sn-ATAC libraries and sn-RNA libraries were simultaneously prepared using Chromium Next GEM Single Cell ATAC Kit v1.1 and Chromium Next GEM Single Cell 3’ Kit v3.1 respectively; following the manufacturer’s instructions. We aimed to recover 10,000 nuclei per sample for both sn-ATAC and sn-RNA libraries. Libraries of the different donors were pooled equimolarly for each of the snATAC and snRNA libraries. Illumina Free Adaptor blocking Reagent was applied as per manufacturer’s instructions. Libraries were sequenced on the NovaSeq 6000 System (Illumina, San Diego, California, USA).

### 4.3. Processing of single-nucleus data

#### 4.3.1. snRNA-seq data workflow

Initial processing of the snRNA-seq data, including the alignment of reads to a pre-mRNA reference (genome build GRCh38, Ensembl 98), cell barcoding and UMI counting, was performed with Cell Ranger (cellranger count v6.0.1)^39^. To account for significant differences in sequencing depth between cells and samples, we downsampled reads to the 75% quantile which corresponds to 14,786 reads per cell. This downsampling procedure was performed with the downsampleReads method from the DropletUtils package v1.12.2^40^, brought the sequencing depth of cells in different samples to a more comparable level and prevented biases in the analysis.

Count matrices of all donors were combined and further processed in Python (Python Software Foundation, https://www.python.org/), primarily using Scanpy v1.7.1^41^. Nuclei were filtered according to counts, minimum genes expressed and percent of mitochondrial genes (counts < 500, genes < 300, Mito % ≥ 15). Genes expressed in < 500 nuclei were removed. One individual was filtered out due to overall low data quality, coinciding with a low RIN value. To ensure data integrity and the accuracy of our analysis, we conducted doublet removal using the DoubletDetection package v3.0^42^. Data was normalized and log-transformed using sctransform v0.3.2^43^. Highly variable genes were identified and dimensionality reduction, including principal component analysis (PCA) and uniform manifold approximation and projection (UMAP) was performed with Scanpy^41^. Nuclei were clustered based on highly variable genes using the leiden clustering algorithm^44^ (resolution 1.0). Four donors were filtered out due to the fact that more than 50% of their nuclei were located within one cluster, resulting in 787,046 nuclei from 87 donors, see Table S2.

#### 4.3.2. snATAC-seq data workflow

The initial processing of the snATAC-seq data, including the alignment of reads to a reference (genome build GRCh38, Ensembl 98), cell calling and count matrix generation, was performed with Cell Ranger ATAC (cellranger-atac count v2.0.0)^45^.

Further processing of the data was performed in R v4.0.5^46^ with the ArchR package v1.0.2^47^. During per-cell quality control, nuclei with a transcription start site (TSS) enrichment score < 4 were excluded due to a low signal-to-noise ratio. Furthermore, nuclei with less than 1,000 or more than 100,000 unique nuclear fragments were filtered out. Doublet scores were inferred in ArchR^47^ and respective doublets were removed with a filter ratio of 2.5. One donor was filtered out due to overall low data quality, coinciding with a low RIN value. Iterative latent semantic indexing (LSI) was used for dimensionality reduction in order to handle the high sparsity of snATAC-seq data. From this lower dimensional space, a UMAP embedding was inferred for visualization purposes. Nuclei were clustered with resolution 1.0 via an interface to the FindClusters method from Seurat v4.0.4^48^ which is based on the louvain clustering algorithm^49^. During a final filtering, another donor with the majority of its nuclei clustering together and six clusters with low data quality regarding doublet scores and number of fragments were removed, resulting in 399,439 nuclei from 90 donors, see Table S2.

To assess chromatin accessibility directly at the gene level, gene scores were calculated with ArchR. Gene scores are estimates of gene expression predicted from the accessibility of the regulatory region surrounding a gene (100 kb up- and downstream), whereby the signal is weighted by the distance to the gene^47^.

#### 4.3.3. Cell type assignment of snRNA-seq and snATAC-seq data

An initial cell type assignment to clusters of nuclei in the snRNA-seq data was carried out using a label transfer algorithm (scArches v0.4.0^50^/scANVI^51^). Thereby, cell type labels were adopted from a cortical dataset of the Allen Brain Map (Human Multiple Cortical Areas^52^) to our snRNA-seq dataset, employing a variational inference model. Each cluster was labeled with the cell type assigned to the majority of nuclei within this cluster. Subsequently, the cell type labels were fine-tuned through manual curation based on the expression of known marker genes, as previously decribed^37^. Marker genes included: Astrocytes: *AQP4*, *CLU*, *GFAP*, *GJA1*; Endothelial: *CLDN5*, *COBLL1*, *FLT1*, *SYNE2*; Excitatory neurons: *SATB2*, *SLC17A6*, *SLC17A7*; Inhibitory neurons: *GAD1*, *GAD2*, *NXPH1*, *SLC32A1*; Microglia: *APBB1IP*, *C3*, *P2RY12*; Oligodendrocytes: *MPB*, *MOBP*, *PLP1*, *RNF220*; Oligodendrocyte Precursors (OPC): *OLIG1*, *OLIG2*, *PDGFRA*, *VCAN*. Astrocyte subtypes: higher *GFAP* and *ARHGEF4* expression (fibrous astrocytes (Astro_FB)) vs. higher expression of *ATP1A2*, *GJA1* and *SGCD* (protoplasmic astrocytes (Astro_PP))^53^. Excitatory neuron subtypes were labeled based on the expression of cortical-layer specific marker genes: layers 2-3: *CUX2*, *RFX3*; layer 4: *IL1RAPL2*, *CRIM1*, *RORB*; layers 5-6: *RXFP1*, *TOX*, *DLC1*, *TLE4*^34,53^. Inhibitory neuron subtypes were labeled based on the expression of interneuron markers *LAMP5*, *PVALB*, *RELN*, *SST*, and *VIP*. PVALB inhibitory neurons consisted of two subtypes: basket cells (In_PVALB_Ba) and chandelier cells (In_PVALB_Ch; identified based on the high expression of *RORA*, *TRPS1*, *NFIB*, and *UNC5B*)^54^.

For the initial assignment of cluster identities in the snATAC-seq data, the data was integrated with snRNA-seq data in ArchR^47^ via a parallelized interface to the FindTransferAnchors function in Seurat^48^. Nuclei from snATAC-seq are getting aligned with nuclei from scRNA-seq by comparing the gene score matrix with the gene expression matrix. Each snATAC-seq nucleus is labeled with the cell type of the most similar scRNA-seq nucleus. Adjacently, cluster identities were refined manually based on gene scores of the marker genes mentioned above. Although known marker genes of endothelial cells did not exhibit distinct gene scores in the cluster labeled as endothelial cells, the cluster’s clear separation of other clusters, the unambiguous assignment as endothelial cells via label transfer and imputed gene scores^55^ allowed for a confident assignment of the cluster as endothelial cells.

#### 4.3.4. Pseudobulk replicates of snRNA-seq and snATAC-seq data

To enable downstream analyses that require replicates with measurements of statistical significance, such as peak calling on ATAC-seq data or differential testing on either data modality, pseudobulk replicates were created. Specifically, gene expression and chromatin accessibility count matrices were summed up from the cells within each cell type-donor pair, creating pseudobulk replicates resembling bulk RNA-seq and ATAC-seq data per cell type. The pseudobulk replicates used for cell type-specific peak calling were generated with an ArchR method summarizing multiple sufficiently similar donors within a cell type to circumvent sparsity. Since such multi-individual pseudobulk replicates are not suitable for our downstream analyses, the ArchR-generated replicates were only used during peak calling.

#### 4.3.5. Peak calling on snATAC-seq data

Peak calling was performed per cell type in ArchR^47^ based on pseudobulk replicates via an interface to MACS2^56^. To facilitate downstream computation, peaks have a fixed width of 501 bp and are merged across pseudobulk replicates and cell types via a ranking of peaks by normalized significance and the iterative removal by overlap. The resulting matrix contains a single merged peak set of fixed-width peaks.

### 4.4. Genotype data

#### 4.4.1. DNA extraction, SNP genotyping and imputation

From 10 mg brain tissue genomic DNA was isolated using the QIAamp DNA mini kit (Qiagen) according to manufacturer’s instructions. Following extraction, DNA samples were concentrated using the DNA Clean & Concentrator-5 (Zymo Research).

Samples were genotyped with Illumina GSA-24v2-0_A1 arrays, following the manufacturer’s protocols (Illumina Inc., San Diego, CA, USA). Quality control (QC) was performed in PLINK v1.90b3.30^57^. Sample QC included removal of donors with a missing rate > 2%, as well as cryptic relatives (PI-HAT > 0.125). Donors with autosomal heterozygosity deviation (|Fhet| > 4 SD) and genetic outliers (distance in ancestry components from the mean > 4 SD) were also excluded. Variants with a call rate < 98%, a minor allele frequency (MAF) < 1%, and p-values from the Hardy-Weinberg equilibrium (HWE) test equal ≤ 10^-6^ were removed during variant QC. Imputation was conducted using shapeit2^58^ and impute2^59^, making use of the 1000 Genomes Phase III reference sample. Imputed SNPs with an INFO score below 0.6, MAF < 1%, or deviation from Hardy-Weinberg equilibrium (p-value < 1 × 10^-5^) were excluded from further analysis, resulting in a final set of 9,652,209 SNPs in 92 donors.

#### 4.4.2. Calculation of polygenic risk scores (PRS)

Summary statistics of GWAS studies for a cross-disorder phenotype^5^, schizophrenia^11^, MDD^12^, bipolar disorder^13^ and height^60^ (as a non-psychiatric control) were used to calculate polygenic risk scores (PRS). Posterior effect sizes were inferred from the GWAS summary statistics using PRS-CS v1.0.0^61^. The linkage disequilibrium (LD) reference panel used was the one based on the European samples of the 1000 Genomes Project phase 3, as accessible on the PRS-CS GitHub page. For schizophrenia, a highly polygenic trait, we set the global shrinkage parameter (phi) of PRS-CS to 0.01, while no specific phi parameter was specified for the other traits, given the larger sample size of the GWAS studies, allowing phi to be derived from the data. PRS per donor were calculated from the previously inferred posterior effect sizes in PLINK v2.00a2.3LM^57^ with the score parameter.

### 4.5. Differential analysis

#### 4.5.1. Definition of disease status for differential testing

Differential expression (DE) and differential accessibility (DA) was tested between all donors with a psychiatric diagnosis (schizophrenia, schizoaffective disorder (SCA), bipolar disorder or MDD) against all donors in the control group. Psychiatric disorders were analyzed as a cross-disorder phenotype due to their shared genetic risk and overlapping symptomatology^3–5^, thereby increasing statistical power of the analyses and enabling the identification of shared molecular dysregulations and underlying pathways.

#### 4.5.2. Definition of groups for testing between high and low genetic risk

To assess DE and DA also with regard to overall genetic predisposition, differential testing was performed between donors with high and low genetic risk for a trait or disease. Acknowledging the consensus within the research community that reliable risk predictions are most feasible at the extreme ends of the PRS distribution^14,62^, we categorized genetic risk into binary groups representing these extremes rather than treating it as a continuum. Specifically, we selected 20 donors with the highest PRS and 20 donors with the lowest PRS within the cohort for each trait or disease.

Subsequently, we employed propensity score matching to identify subsets of extreme groups that are matched based on key covariates such as age, sex, brain pH, PMI, RIN (Figure 4a). Sex was matched exactly (Supp. Figure 6). We used the matchit function from MatchIt v4.5.5^63^ for this purpose. The resulting number of donors in each extreme group ranges from 11 to 17, with specific counts being: 17 donors for both high and low PRS groups in cross-disorder, 13 donors for both groups in schizophrenia, 14 donors for both groups in bipolar disorder, 11 donors for both groups in MDD, and 14 donors for both groups in height.

#### 4.5.3. Selection of covariates for differential testing between disease status and extreme genetic risk groups

To comprehensively evaluate the impact of biological variables and batch effects on the data and to select relevant covariates for differential testing, we assessed the impact of potential confounders on the RNA-seq data. Given the assumption that technical covariates remain consistent across cell types, a full pseudobulk count matrix was created by summing gene-wise counts across all cell types. Only genes with a minimum of 10 counts in at least 90% of the samples were retained for the covariate selection process. Data was normalized with the variance stabilizing transformation in DESeq2^64^ and principal component analysis was applied. A significant correlation between continuous variables and one of the first 10 principal components was observed for RNA integrity number (RIN), postmortem interval (PMI), pH and age. Further exploration using canonical correlation analysis identified the library preparation batch (lib_batch) as a covariate. However, the inclusion of the library preparation batch into the model was limited to disease status, owing to the insufficiency of observations within each batch in the genetic risk model to support a categorical variable in the genetic risk model. Additionally, we included sex as a commonly known confounder as a covariate into our model.

To account for hidden noise, principal component analysis was performed after having normalized and transformed the data and regressed out the effect of all mentioned covariates and our variable of interest (disease status (Disease_Status) or genetic risk group (Genetic_Risk) respectively) of the data using voom and removeBatchEffect from the limma package v3.56^65^. We included the first principal component (PC_noise) as additional covariate into our final model for differential testing: (∼Disease_Status/Genetic_Risk + Sex + Age + pH + RIN + PMI + lib_batch + PC_noise). RIN was not present for one donor and therefore imputed to the median value across the cohort.

To keep analyses consistent and due to the fact that snRNA-seq and snATAC-seq data were generated from the exact same tissue, and library preparation was performed in the same batches for both data modalities, the same covariates – except RIN – were included into the final model of differential chromatin accessibility analysis.

#### 4.5.4. Differential expression analysis

DE was tested on the pseudobulk level with DESeq2 v1.40.2^64^. For each cell type-specific count matrix, genes were filtered for a minimum of 10 counts in 75% of the pseudobulk samples. After data normalization with the variance stabilizing transformation in DESeq2^64^, outlier samples were excluded by iterative PCA and the removal of samples with a distance of more than 3 standard deviations from the mean on the first PC. We tested for DE with DESeq2 using the Wald test. Genes with a false discovery rate (FDR) ≤ 10% were reported as significant, given that the pseudobulk approach is considered more conservative than single-cell DE methods^66,67^.

#### 4.5.5. Differential chromatin accessibility analysis

DA was tested on the pseudobulk level for each cell type using gene scores. As gene scores do not follow the typical characteristics of count data, differential testing was not performed with DESeq2. Pseudobulk gene scores were normalized by the number of cells aggregated per pseudobulk sample and outliers were filtered the same way as during DE analysis. Genes exhibiting scores above 0.1 in less than 75% of the samples were filtered out and removed from further analysis. After the fitting of a linear model including the previously described covariates, a Wald test was performed and log_2_-fold changes were calculated. Genes with FDR ≤ 10% were considered as significant.

#### 4.5.6. Differential risk group analysis

Differential risk group analyses of gene expression (DE risk analysis) and chromatin accessibility (DA risk analysis), comparing donors in high and low genetic risk groups for a phenotype, was performed analogously to differential testing between cases and controls. Gene filtering, normalization and outlier removal followed the same principles and genes with FDR ≤ 10% were considered significant.

### 4.6. Functional annotation

#### 4.6.1. Pathway enrichment analysis

Pathway enrichment analysis was conducted with clusterProfiler v4.8.1^68^. The 250 genes with the most significant up- and downregulation for each cell type according to FDR values were assessed for over-representation of KEGG pathways. The choice of not only testing the significant DE and DA genes was made in order to make this analysis comparable between cell types. 250 genes per direction of downregulation corresponds to about 50% of the number of DE genes in Exc_L2-3, the cell type with the highest number of DE genes between cases and controls. Any KEGG pathway significant in at least one cell type (FDR ≤ 0.05) is shown in the respective heatmaps. To summarize single KEGG pathways in categories, a hierarchy of KEGG pathways was downloaded from the KEGG Pathway Database (https://www.genome.jp/kegg/pathway.html, accessed: June 19th, 2023) and used to annotate the enrichment heatmap.

#### 4.6.2. Transcription factor motif enrichment analysis

To assess if peaks in the promoter regions of a given gene are enriched for binding sites of specific transcription factors (TFs), a TF motif enrichment analysis was performed within the ArchR framework. As an initial step, the addMotifAnnotation function was used to obtain binary information for each peak-TF pair whether a respective motif is present in the peak or not. TF motif information was obtained from the JASPAR 2020 database^69^. Subsequently, an adapted version of the peakAnnoEnrichment was applied to test the peaks in a given gene’s promoter region for enriched presence of TF motifs compared to the presence in all peaks using a hypergeometric test. TF motifs with an adjusted p-value ≤ 0.05 were reported as significantly enriched.

### 4.7. Comparison to previous findings

We conducted a comparative analysis of DE results for disease status against previously documented cell type-specific transcriptomic changes in the prefrontal cortex of schizophrenia patients. This comparison aimed to evaluate the reproducibility of our findings in relation to other studies. Effect sizes were correlated with those reported in a single-cell RNA-seq meta-analysis by Ruzicka et al. (sample sizes: 140; cell counts: 469K)^26^. For each cell type pair, we calculated Pearson’s correlation coefficient to measure the relationship between effect sizes for all genes examined in both studies.

### 4.8. Network inference

For given genes which are differentially expressed and accessible between extreme genetic risk groups for schizophrenia, correlation-based networks were inferred to integrate gene expression and chromatin accessibility across the different cell types as well as disease status and PRS for the aforementioned disorders and traits. The analysis was based on the donors that are part of the extreme genetic risk groups for schizophrenia. Gene expression and gene score levels were normalized and corrected for sex, age, RIN, PMI, pH and the library preparation batch. Spearman correlation was calculated between each pair of features. Correlations with a nominal p-value ≤ 0.05 are shown in each network and edge strength/weight transfers to the absolute correlation coefficient. Networks were visualized with the igraph^70^ and ggnetwork^71^ packages in R.

## Acknowledgements

This work was supported by the Hope for Depression Research Foundation. Human brain tissue acquisition was funded by the Alexander von Humboldt Foundation research support package awarded to Dr. Natalie Matosin. Nathalie Gerstner is supported by the Joachim Herz Foundation. The authors would like to thank Dr. Darina Czamara and Dr. Jade Martins for processing genotype data and performing SNP imputation, Dr. Simone Roeh and Vanessa Murek for their help in preprocessing the sequencing data and Susann Sauer for her support in prepping samples for genotyping. We thank the Medical Genomics Group and the Translational Genomics Team (Max Planck Institute of Psychiatry) for the useful discussions during the project implementation. The authors would also like to thank the donors and their families for the donations of brain tissue, as well as the staff at the New South Wales Brain Tissue Resource Centre for tissue collection and processing. Tissues received from the New South Wales Brain Tissue Resource Centre at the University of Sydney were supported by the University of Sydney. Research reported in this publication was supported by the National Institute of Alcohol Abuse and Alcoholism of the National Institutes of Health under Award Number NIAAA012725-15. The content is solely the responsibility of the authors and does not represent the official views of the National Institutes of Health.

## Author contributions

E.B.B., A.S.F, J.K.A., N.G. and M.J.Z. conceptualized single-cell study design. J.K.A. and N.G. equally contributed to the analysis strategy design, result interpretation and writing of the manuscript. J.K.A designed and supervised the study. N.G. performed read alignment, quality control of single-nuclei data, label transfer for cell type assignment, all computational analyses and data visualization. A.S.F. performed DNA extraction and manual cell type label curation. A.S.F., M.G. and M.K. performed single-nuclei extraction and library preparation. M.K. prepped samples for genotyping. M.R.H. contributed to project organization and logistics. X.T. and S.M. provided intellectual input for the analysis of the single-nuclei ATAC-seq data. N.M. and C.C. provided intellectual input and performed critical revisions of the manuscript. E.B.B. and M.J.Z. obtained funding for the study. N.M. curated the cohort, acquired, managed, and funded the human brain tissue acquisition. All authors reviewed and revised the manuscript.

## Data availability

Significant results from differential analysis of gene expression and chromatin accessibility between cases and controls as well as high and low risk individuals are available in Table S4-S9. Complete result lists can be obtained from https://doi.org/10.5281/zenodo.10522979. Preprocessed and raw single-nuclei data are available upon reasonable request to the authors.

## Code availability

Data analysis scripts and scripts that were used to generate the manuscript figures are available via github: https://github.molgen.mpg.de/mpip/SingleNuclei_Analysis_OFC.git

