## Supplemental Material for "Contrasting genetic predisposition and diagnosis in psychiatric disorders: a multi-omic single-nucleus analysis of the human orbitofrontal cortex"

### Supplemental Information

**Supplemental Methods** **1**
1. Correlation analysis between gene expression and chromatin accessibility 1
2. Downsampling of nuclei 2
3. GWAS enrichment analysis 2

**Supplemental Results** **2**
1. Cell type-specific cis-regulatory gene regulation 2
2. Differences in detection power between cell types 3
3. GWAS enrichment analysis using H-MAGMA 3

**Supplemental Figures** **4**

### Supplemental Methods

#### 1. Correlation analysis between gene expression and chromatin accessibility

The assessment of the number of peaks nearby each gene and the number of nearby correlated peaks per gene was performed for each cell type separately on the pseudobulk level. Peaks with less than 5 counts in more than 50% of the samples were removed from the peak matrix and genes with less than 5 counts in more than 75% were excluded from the count matrix. The less stringent filtering in the peaks was applied due to the even sparser signal in ATAC-seq data. Gene expression and peak matrix were normalized with the variance stabilizing transformation in DESeq2[64]. Peaks within a 100 kb window from the gene body, the default distance used to calculate gene scores in ArchR[47], were considered to be nearby a gene and tested for correlation. Pearson's correlation coefficient was used as a measure of the association between gene expression and chromatin accessibility.

In addition to the correlation between gene expression and chromatin accessibility on the peak level, expression levels were also correlated with gene scores. This analysis was performed in each cell type separately and across all cell types. On the pseudobulk level, Pearson's correlation coefficients were calculated across all genes between expression and gene scores averaged across all samples. Additionally, the distribution of Pearson's correlation coefficients calculated between gene expression and gene scores across all pseudobulk samples for each gene was compared to a random distribution obtained by correlating gene expression with a random permutation of gene scores.

#### 2. Downsampling of nuclei

To dissect to what extent the number of DE genes in a cell type is influenced by its nuclei count and consequently the number of genes tested for differential expression, a downsampling analysis was performed. The nuclei per cell type were downsampled to the 75%, 50% and 25% percentiles of nuclei (40,793, 31,504, 14,416 nuclei respectively). Differential expression analysis was performed on the downsampled datasets as described in Methods.

#### 3. GWAS enrichment analysis

GWAS enrichment analysis was performed with H-MAGMA v1.10[110]. A mapping of SNPs to genes was generated based on GWAS summary statistics for schizophrenia[11], bipolar disorder[13] and MDD[12] and the european 1,000 genomes reference panel downloaded from the H-MAGMA github page (https://github.com/thewonlab/H-MAGMA). Based on these results, a gene-level analysis in the form of a gene property analysis was performed with the "--gene-covar" argument in MAGMA. This analysis allows the input of a continuous variable (here: DE (risk) results in the form of $-\log_{10}$(p-value)*$\log_{2}$(fold change)) into the gene-level regression framework to test if DE related to disease status/genetic risk is associated with GWAS results.

### Supplemental Results

#### 1. Cell type-specific cis-regulatory gene regulation

To elucidate the specific cis-regulatory interactions between chromatin accessibility and gene expression within distinct cell types, independent of any disease phenotype influence, we conducted a thorough analysis. This involved quantifying the number of proximate peaks (within 100 kb of the gene body) for each gene. Subsequently, we correlated the signal of these peaks with gene expression levels after applying appropriate filtering and normalization techniques specific to each cell type.

For example, in oligodendrocyte precursor cells (OPCs), the range of nearby peaks within the 100 kb region surrounding the gene body ranges from 0 to 112, with a median count of 6. More than 1,500 genes exhibited no proximate peaks in their vicinity (Supp. Figure 2a). While similar patterns emerged for other cell types, they displayed distinct maximum values, consistently low median counts, and a substantial number of genes lacking nearby peaks within the 100 kb region from the gene body. Among the peaks situated near a gene, even fewer demonstrated a significant correlation with the respective gene's expression levels. The maximum number of peaks showing nominal significance (p-value $\leq 0.05$) was 18, while nearly 8,000 genes were without any correlated peaks (Supp. Figure 2b).

Due to this sparse signal on the peak level, we examined the relationship between gene expression and chromatin accessibility on the gene level, making use of gene scores which predict the level of gene expression from the accessibility of gene regulatory elements nearby a gene without the necessity to call peaks. The correlation between the mean normalized gene expression values and gene scores across donors significantly correlates across ($R = 0.47$, Supp. Figure 2c) and within cell types ($R = [0.4, 0.56]$ in all cell types, Supp. Figure 2d). While the correlation between normalized gene expression and gene scores remains high if we correlate the respective pseudobulk samples across all cell types (Supp. Figure 2e), thereby keeping cell type and sample-specific differences in the data, correlations are rather low and partly even negative if we correlate only pseudobulk samples within a specific cell type, thereby keeping only sample-specific differences (Supp. Figure 2f). However, the distribution of correlations is still significantly different from a random distribution, generated with a permutation of the gene scores across pseudobulk samples.

As a result, we opted to conduct downstream analyses at the gene score level. This approach addresses the challenge of missing (and correlated) peaks for numerous genes, providing a more comprehensive view of the regulatory landscape surrounding each gene.

### 2. Differences in detection power between cell types

The number of DE genes in each cell type was influenced by its nuclei count (Supp. Figure 3a) and consequently the number of genes tested. Downsampling the nuclei per cell type to the 75%, 50% and 25% percentiles of nuclei (n=40,793, 31,504, 14,416 respectively), revealed that the gap between the number of tested genes and DE genes in excitatory neurons and other cell types becomes smaller with the level of downsampling, but excitatory neurons still exhibited the highest number of DE genes (Supp. Figure 3b-c).

### 3. GWAS enrichment analysis using H-MAGMA

To ascertain whether DE risk genes for specific traits and cell types are enriched for GWAS-associated genes for psychiatric disorders (bipolar disorder[13], MDD[12] and schizophrenia[11]), we conducted a GWAS enrichment analysis using H-MAGMA[110]. No significant enrichments of GWAS-associated genes emerged among the DE risk results for cross-disorder phenotype, bipolar disorder, MDD and height. However, we identified significant enrichments of schizophrenia GWAS-associated genes in the DE risk results for schizophrenia within basket cells (In_PVALB_Ba), excitatory neurons layers 2 to 3 (Exc_L2-3) and endothelial cells (Figure 7b). Similarly, MDD GWAS-associated genes exhibited significant enrichment in endothelial cells' DE risk results for schizophrenia (Figure 7b).

### Supplemental Figures

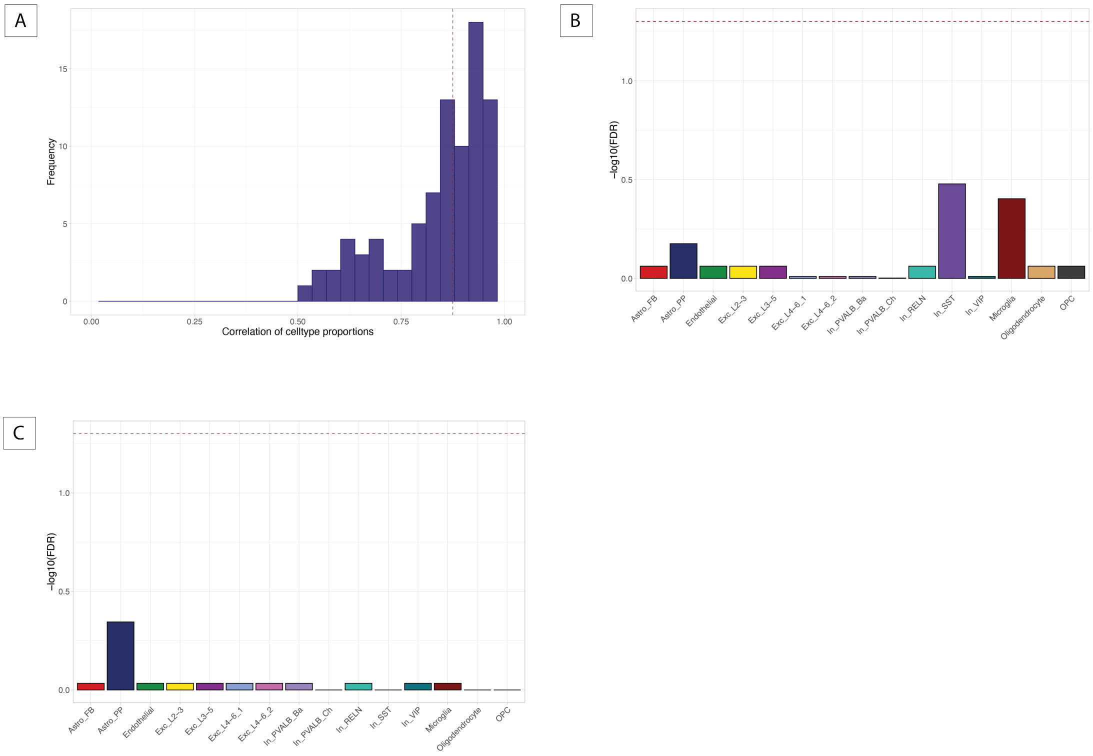


**Supp. Figure 1. Differences in cell type proportions between data modalities and disease status.** (A) Histogram of Pearson correlation coefficients between cell type proportions in snRNA-seq and snATAC-seq data across all donors. (B-C) Significance of differences in cell type proportions between snRNA-seq cases and controls (B) and snATAC-seq cases and controls (C). Height of the bar represents -$log_{10}$-transformed FDR values of Wilcoxon rank-sum test and the dashed red line corresponds to the FDR cutoff of 0.05.

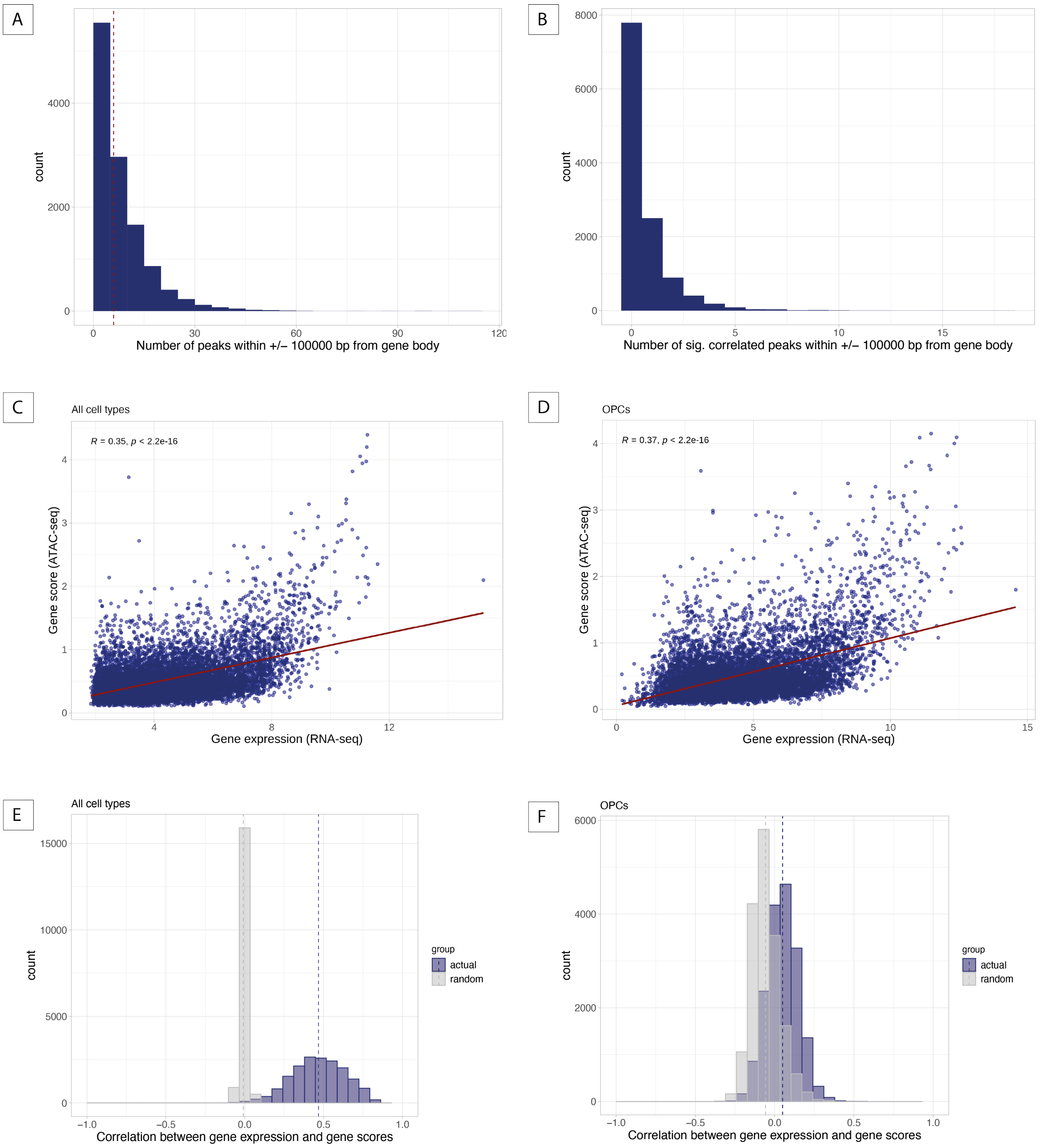


**Supp. Figure 2. Cis-regulatory interactions between chromatin accessibility and gene expression.** OPCs are chosen as an exemplary cell type in this figure. (A) Histogram of the number of peaks within a 100kb window from the gene body for all genes tested for differential expression in OPCs. Dashed red line indicates the median number of peaks. (B) Histogram of the number of nominally significantly correlated peaks ($P \leq 0.05$) within a 100kb window from the gene body in OPCs. (C-D) Mean gene expression levels plotted against mean gene score levels across all cell types (C) and in OPCs (D). The red line represents a linear model fitted on the data. Pearson correlation is shown in the upper left corner. (E-F) Histogram of correlations between gene expression and gene score levels on the donor level across all cell types (E) and in OPCs (F). The distribution of the actual correlation coefficients (blue) is plotted along the distribution obtained by randomly permuting the donor levels (gray). The dashed line indicates the mean values respectively.

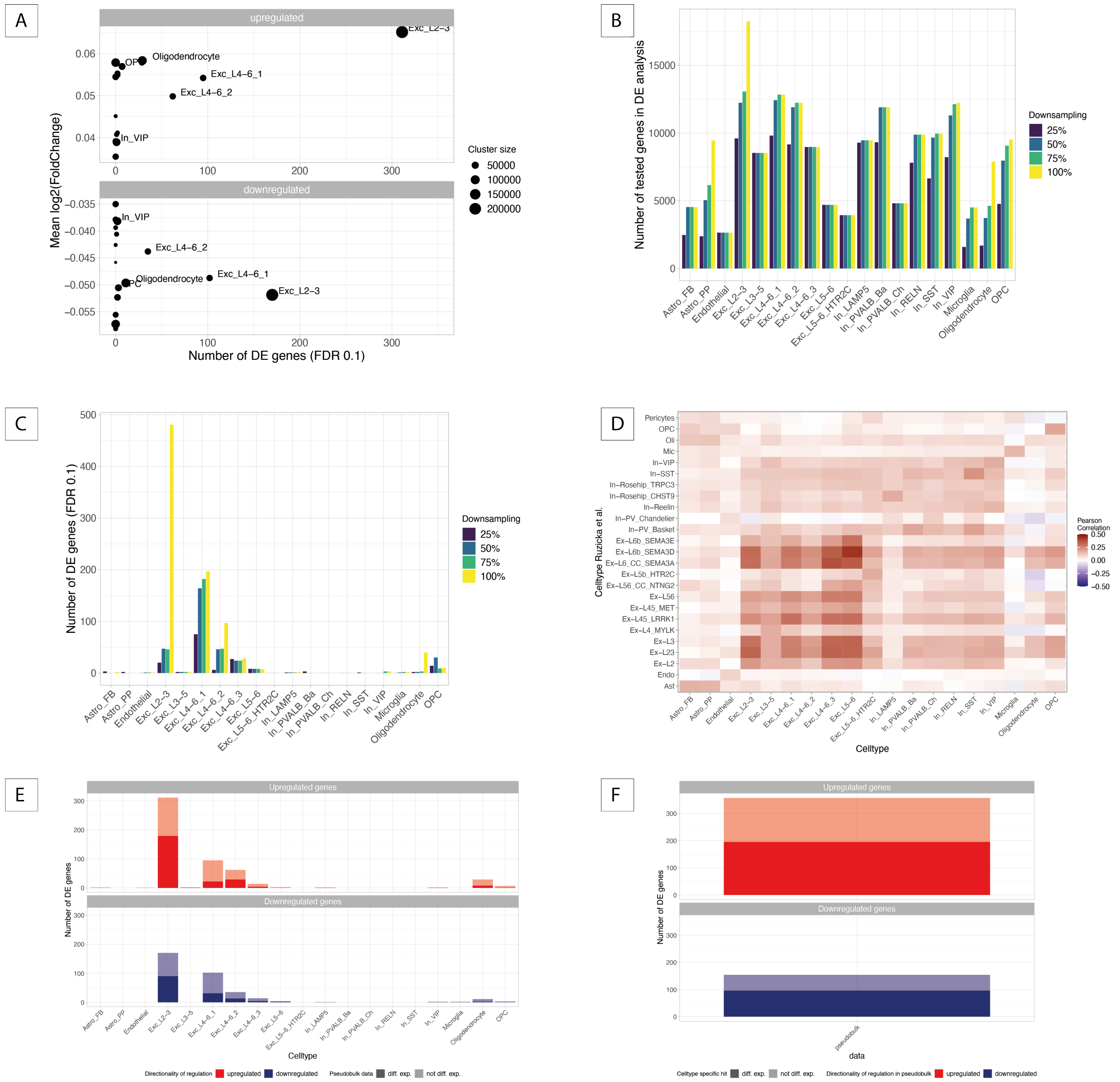


**Supp. Figure 3. Transcriptional alterations between psychiatric cases and controls.** (A) Number of DE genes (FDR ≤ 0.1) plotted against the mean $log_2$-fold change for up- and downregulated genes separately. Dot size indicates the cluster size. (B-C) Barplot representing the number of genes tested for differential expression (B) and the number of significant DE genes (C) using the full dataset and datasets downsampled to the 75%, 50% and 25% percentile of nuclei per cell type which is indicated by color. (D) Correlation analysis between effect sizes from our study and effect sizes reported in Ruzicka et al.[26] for each pair of cell types. Color indicates the Pearson correlation coefficient based on the effect sizes for the shared set of genes tested in the respective cell types. (E) Barplot representing the number of DE genes per cell type for up- and downregulation separately and the proportion of genes also identified as DE based on full pseudobulk data, represented by darker color. (F) Barplot representing the number of DE genes based on full pseudobulk data for up- and downregulation separately. The darker parts of the bars represent the proportion of genes also identified as DE in at least one cell type.

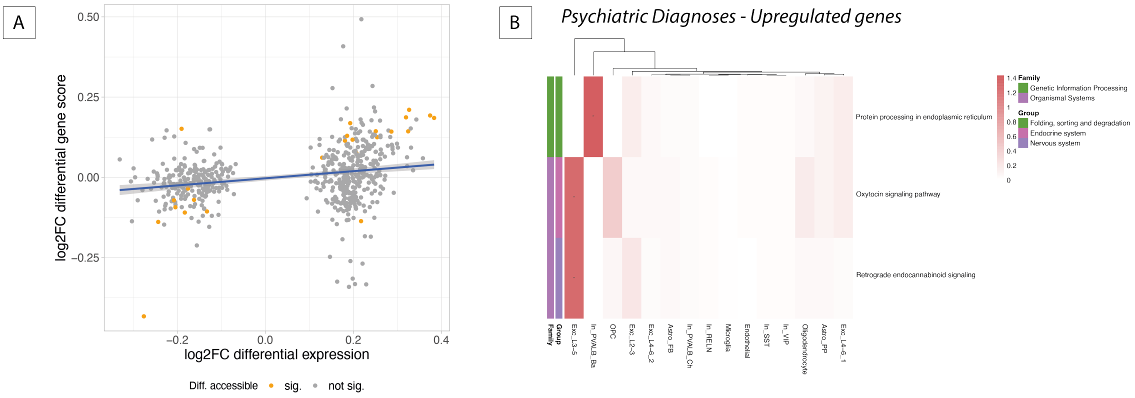


**Supp. Figure 4. Epigenomic alterations between psychiatric cases and controls.** (A) $Log_2$-fold changes of differential expression and accessibility analysis for all DE genes across cell types plotted against each other with significance in the same cell type indicated by color. The blue line represents a linear model fitted on the data. (B) Results of KEGG pathway enrichment analysis for 250 most up- and downregulated genes per cell type. All pathways significantly enriched in at least one cell type are included into the heatmap. Color represents -log10-transformed FDR values and asterisks indicate significance (FDR $\leq$ 0.05). Colored annotations of the pathways on the left side of each plot indicate to which pathway group and family a pathway belongs. The dendrograms visualize k-means clustering of cell types according to enrichment results.

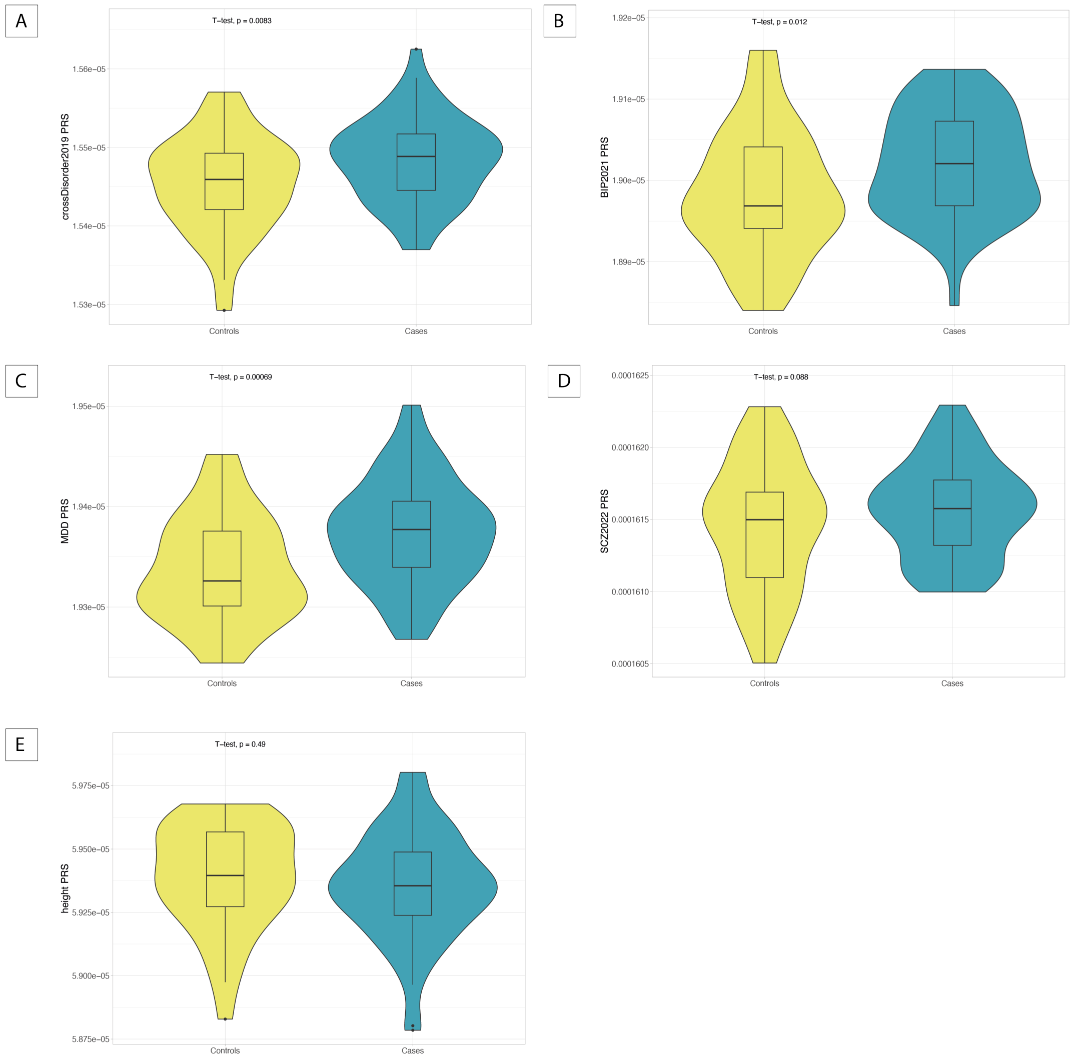


**Supp. Figure 5. Genetic risk for psychiatric disorders in cases and controls.** (A-E) Distribution of polygenic risk scores (PRS) for cross-disorder phenotype (A), bipolar disorder (B), MDD (C), schizophrenia (D) and height (E) for controls and cases. A one-sided t-test was used to test for differences in cross-disorder, bipolar disorder, MDD and schizophrenia PRS between cases and controls, while a two-sided t-test was used to test for differences in height PRS.

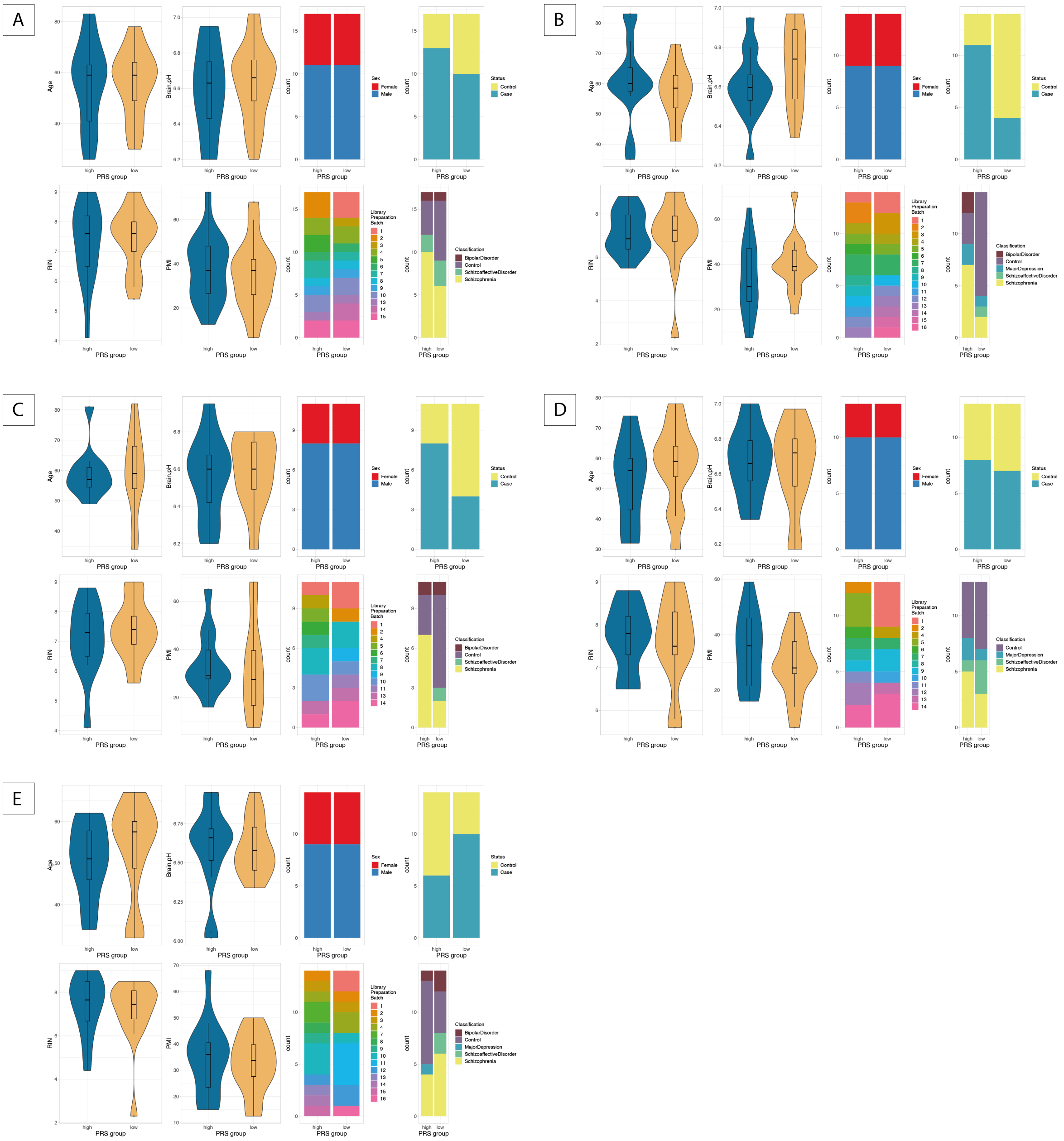


**Supp. Figure 6. Definition of extreme groups for genetic risk.** (A-E) Matched covariates and distribution of disease status and diagnoses for high and low risk groups for cross-disorder phenotype (A), bipolar disorder (B), major depressive disorder (C), schizophrenia (D) and height (E).

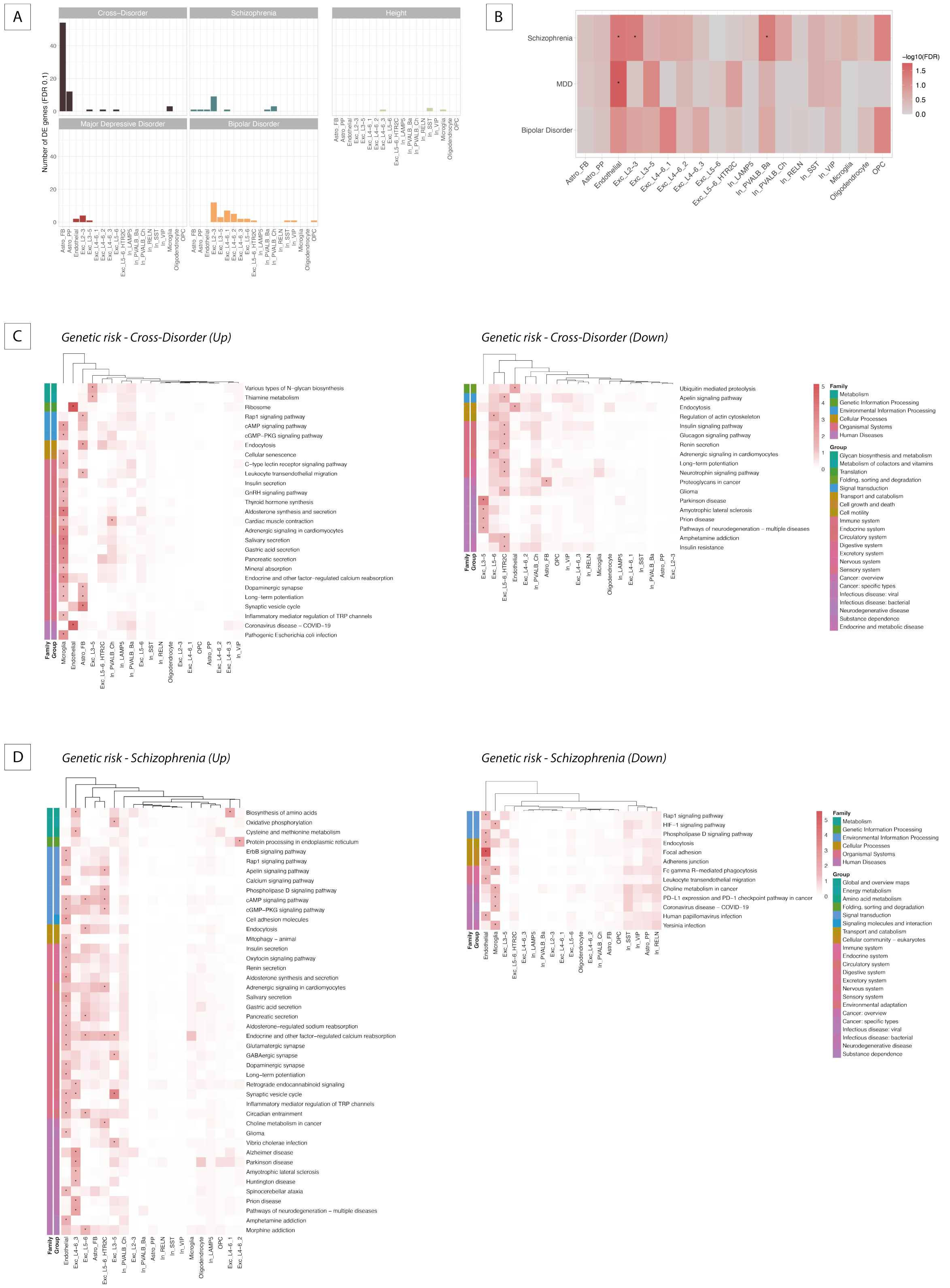
A
Cross−Disorder
Schizophrenia
Height
Major Depressive Disorder
Bipolar Disorder
Number of DE genes (FDR 0.1)
B
Schizophrenia
MDD
Bipolar Disorder
-log10(FDR)
C
Genetic risk - Cross-Disorder (Up)
Genetic risk - Cross-Disorder (Down)
Various types of N−glycan biosynthesis
Thiamine metabolism
Ribosome
Rap1 signaling pathway
cAMP signaling pathway
cGMP−PKG signaling pathway
Endocytosis
Cellular senescence
C−type lectin receptor signaling pathway
Leukocyte transendothelial migration
Insulin secretion
GnRH signaling pathway
Thyroid hormone synthesis
Aldosterone synthesis and secretion
Cardiac muscle contraction
Adrenergic signaling in cardiomyocytes
Salivary secretion
Gastric acid secretion
Pancreatic secretion
Mineral absorption
Endocrine and other factor−regulated calcium reabsorption
Dopaminergic synapse
Long−term potentiation
Synaptic vesicle cycle
Inflammatory mediator regulation of TRP channels
Coronavirus disease − COVID−19
Pathogenic Escherichia coli infection
Ubiquitin mediated proteolysis
Apelin signaling pathway
Endocytosis
Regulation of actin cytoskeleton
Insulin signaling pathway
Glucagon signaling pathway
Renin secretion
Adrenergic signaling in cardiomyocytes
Long−term potentiation
Neurotrophin signaling pathway
Proteoglycans in cancer
Glioma
Parkinson disease
Amyotrophic lateral sclerosis
Prion disease
Pathways of neurodegeneration − multiple diseases
Amphetamine addiction
Insulin resistance
Family
Group
D
Genetic risk - Schizophrenia (Up)
Genetic risk - Schizophrenia (Down)
Biosynthesis of amino acids
Oxidative phosphorylation
Cysteine and methionine metabolism
Protein processing in endoplasmic reticulum
ErbB signaling pathway
Rap1 signaling pathway
Apelin signaling pathway
Calcium signaling pathway
Phospholipase D signaling pathway
cAMP signaling pathway
cGMP−PKG signaling pathway
Cell adhesion molecules
Endocytosis
Mitophagy − animal
Insulin secretion
Oxytocin signaling pathway
Renin secretion
Aldosterone synthesis and secretion
Adrenergic signaling in cardiomyocytes
Salivary secretion
Gastric acid secretion
Pancreatic secretion
Aldosterone−regulated sodium reabsorption
Endocrine and other factor−regulated calcium reabsorption
Glutamatergic synapse
GABAergic synapse
Dopaminergic synapse
Long−term potentiation
Retrograde endocannabinoid signaling
Synaptic vesicle cycle
Inflammatory mediator regulation of TRP channels
Circadian entrainment
Choline metabolism in cancer
Glioma
Vibrio cholerae infection
Alzheimer disease
Parkinson disease
Amyotrophic lateral sclerosis
Huntington disease
Spinocerebellar ataxia
Prion disease
Pathways of neurodegeneration − multiple diseases
Amphetamine addiction
Morphine addiction
Rap1 signaling pathway
HIF−1 signaling pathway
Phospholipase D signaling pathway
Endocytosis
Focal adhesion
Adherens junction
Fc gamma R−mediated phagocytosis
Leukocyte transendothelial migration
Choline metabolism in cancer
PD−L1 expression and PD−1 checkpoint pathway in cancer
Coronavirus disease − COVID−19
Human papillomavirus infection
Yersinia infection
Family
Group

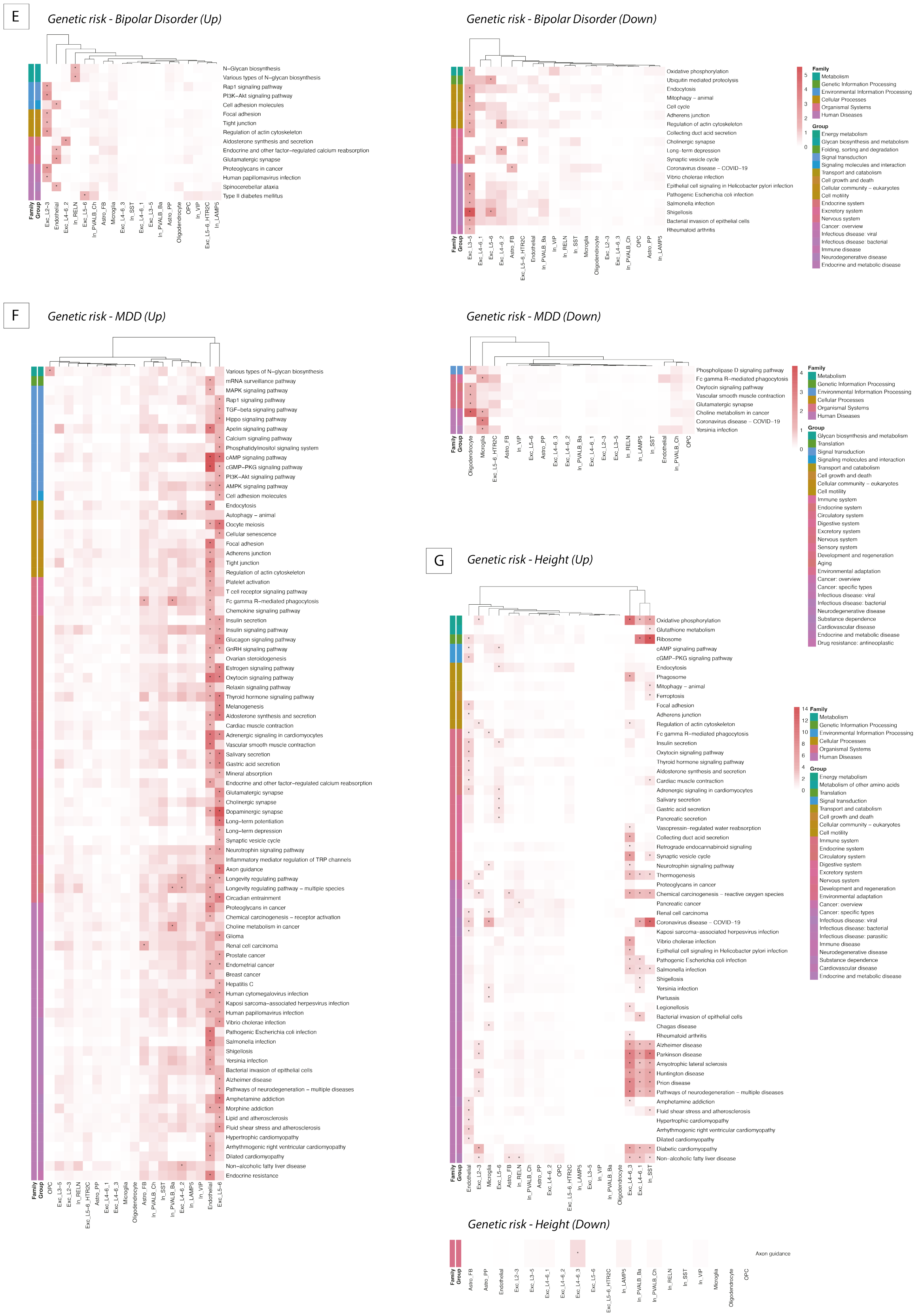


**Supp. Figure 7. Dysregulations in gene expression between extreme genetic risk groups.** (A) Barplot visualizing the number of DE risk genes (FDR ≤ 0.1) between donors at high and low genetic risk quantified by PRS based on 5 different GWAS studies. (B) Results of GWAS enrichment analysis in schizophrenia DE risk genes using H-MAGMA[110] for GWAS hits of bipolar disorder, major depressive disorder and schizophrenia. Color indicates -$\log_{10}$-transformed FDR values and asterisks indicate significance ($FDR \leq 0.05$). (C-G) Results of KEGG pathway enrichment analysis for 250 most up- and downregulated genes per cell type between extreme genetic risk groups for cross-disorder phenotype (C), schizophrenia (D), bipolar disorder (E), MDD (F), and height (G). Left heatmap of each panel shows enrichment results for upregulated genes, while right heatmap of each panel shows results for downregulated genes. All pathways significantly enriched in at least one cell type are included into the heatmap. Color represents -$\log_{10}$-transformed FDR values and asterisks indicate significance ($FDR \leq 0.05$). Colored annotations of the pathways on the left side of each plot indicate to which pathway group and family a pathway belongs. The dendrograms visualize k-means clustering of cell types according to enrichment results.

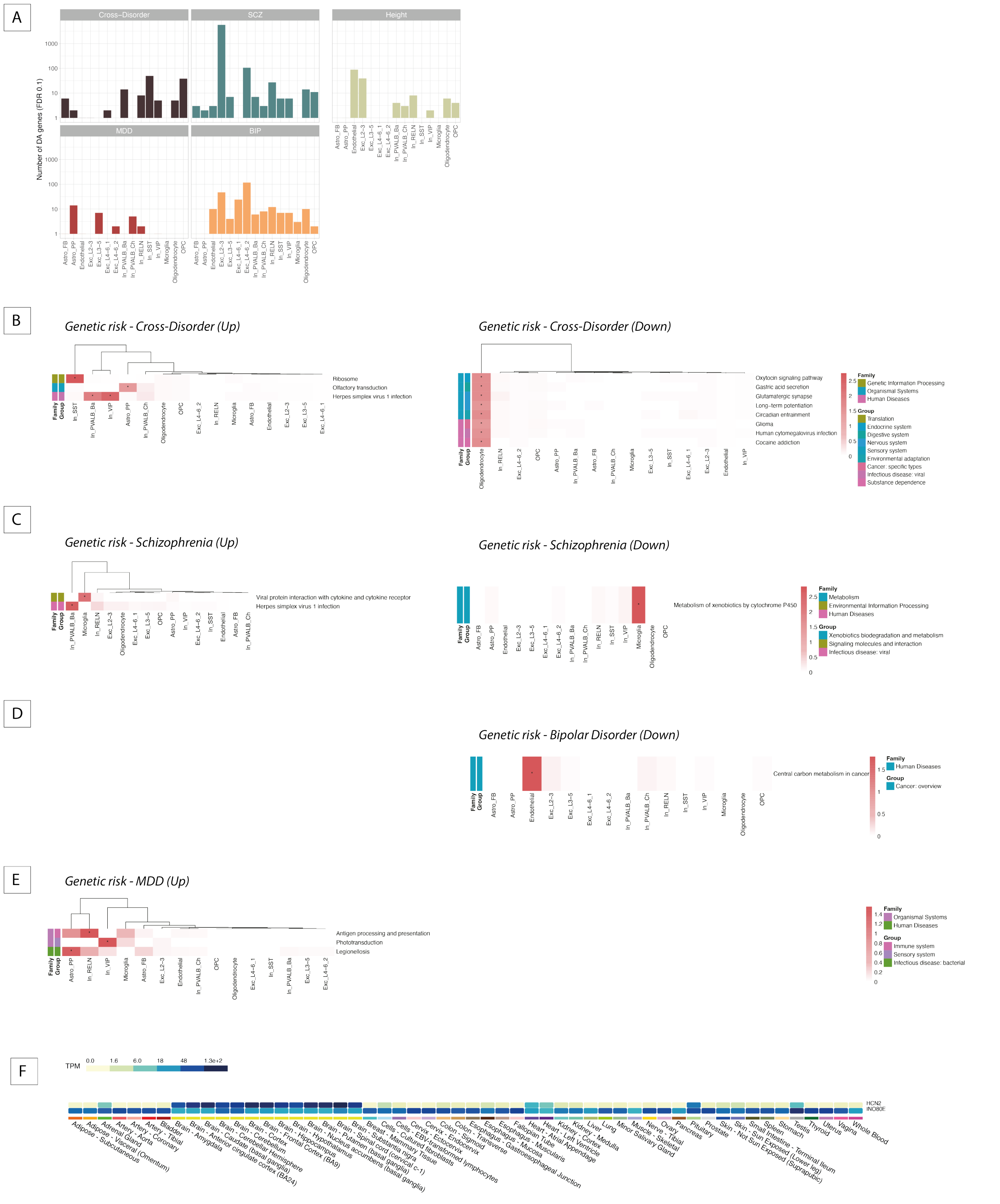


**Supp. Figure 8. Dysregulations in chromatin accessibility between extreme genetic risk groups.** (A) Barplot visualizing the number of DA risk genes (FDR ≤ 0.1) between donors at high and low genetic risk quantified by PRS based on 5 different GWAS studies. (B-E) Results of KEGG pathway enrichment analysis for 250 most up- and downregulated genes per cell type between extreme genetic risk groups for cross-disorder phenotype (B), schizophrenia (C), bipolar disorder (D), and MDD (F). No pathways were enriched for height DA risk genes. Left heatmap of each panel shows enrichment results for upregulated genes, while right heatmap of each panel shows results for downregulated genes. All pathways significantly enriched in at least one cell type are included into the heatmap. Color represents -log$_{10}$-transformed FDR values and asterisks indicate significance (FDR ≤ 0.05). Colored annotations of the pathways on the left side of each plot indicate to which pathway group and family a pathway belongs. The dendrograms visualize k-means clustering of cell types according to enrichment results. (F) Gene expression levels (i.e. transcript per million, TPM) for *HCN2* and *INO80E* across multiple human tissues, as sourced from the GTEx portal[75].

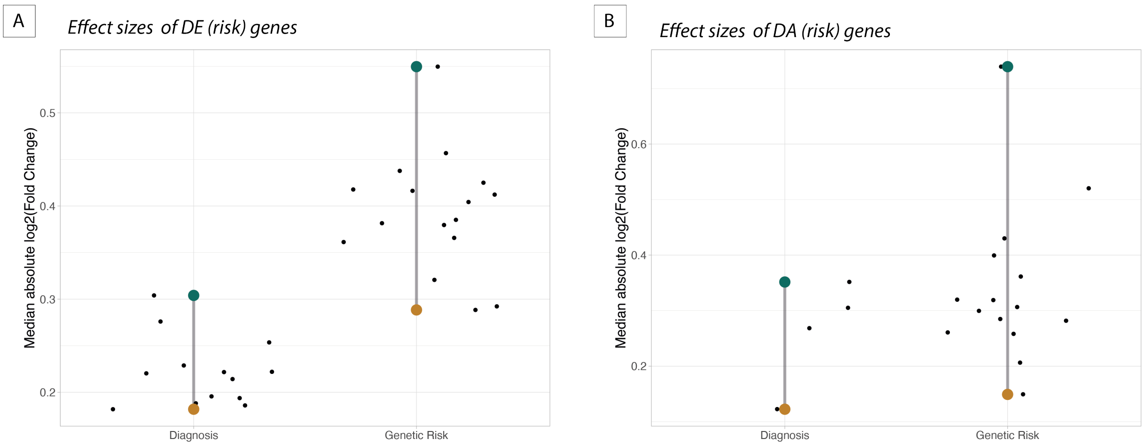


**Supp. Figure 9. Range of effect sizes for clinical diagnosis and genetic risk.** (A-B) Visualizations of the range of absolute median $\log_2$-transformed fold changes per cell type for DE (risk) genes (A) and DA (risk) genes (B). The vertical lines represent the range of effect sizes across cell types with the colored dots representing the minimum and maximum effect size each. Small black dots represent the median effect sizes for specific cell types.

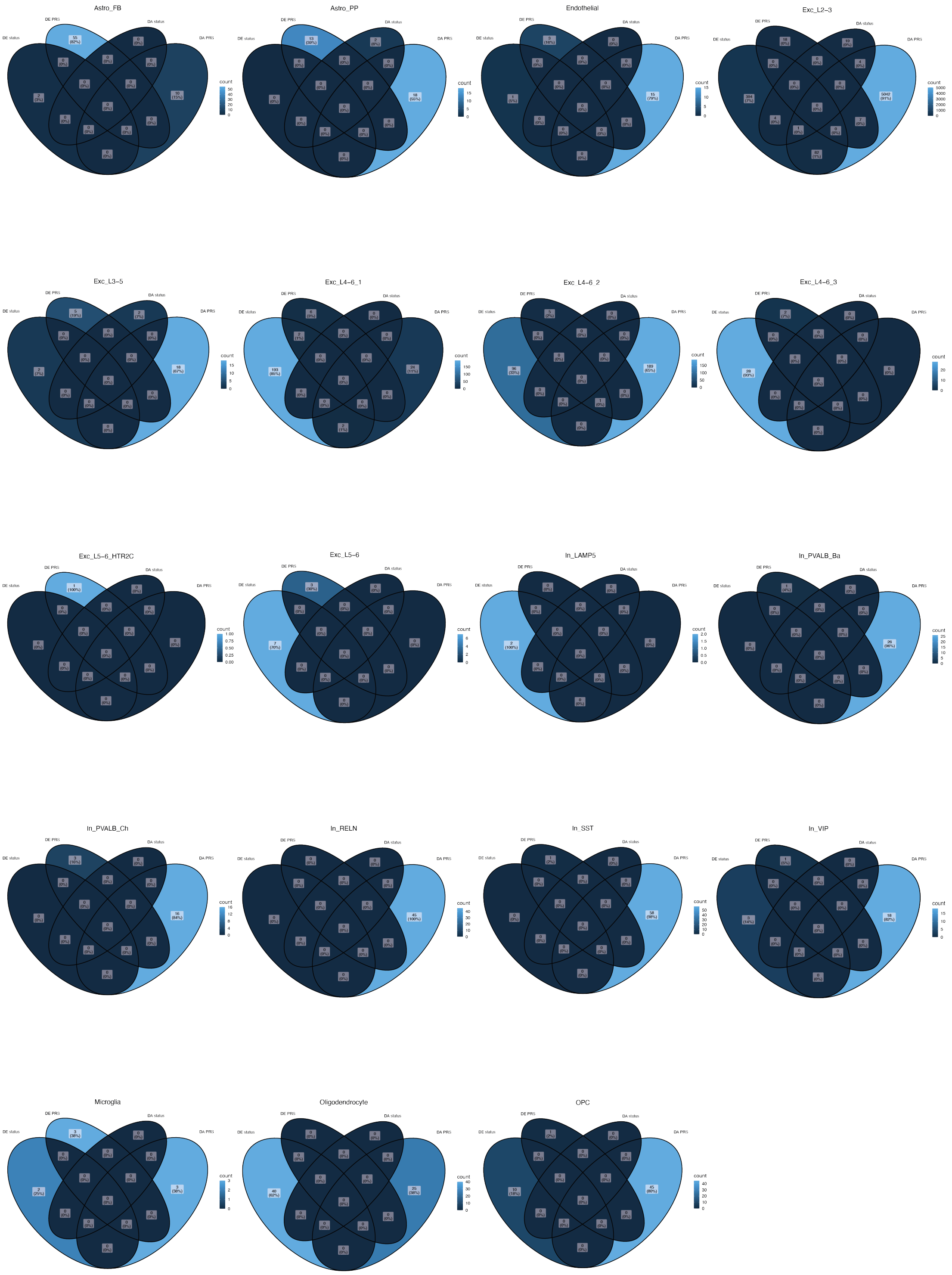


**Supp. Figure 10. Differentially expressed and accessible genes between cases and controls and genetic risk groups.** Venn diagram for each cell type comparing DE and DA genes between disease status with DE and DA risk genes between genetic risk groups. DE and DA risk genes are aggregated across the cross-disorder, schizophrenia, MDD and bipolar disorder GWAS studies.
